# Broad immunity to SARS-CoV-2 variants of concern mediated by a SARS-CoV-2 receptor-binding domain protein vaccine

**DOI:** 10.1101/2022.08.05.22278425

**Authors:** Georgia Deliyannis, Nicholas A. Gherardin, Chinn Yi Wong, Samantha L. Grimley, James P. Cooney, Samuel Redmond, Paula Ellenberg, Kathryn Davidson, Francesca L. Mordant, Tim Smith, Marianne Gillard, Ester Lopez, Julie McAuley, Chee Wah Tan, Jing Wang, Weiguang Zeng, Mason Littlejohn, Runhong Zhou, Jasper Fuk-Woo Chan, Zhi-wei Chen, Airn E. Hartwig, Richard Bowen, Jason M. Mackenzie, Elizabeth Vincan, Joseph Torresi, Katherine Kedzierska, Colin W. Pouton, Tom Gordon, Lin-fa Wang, Stephen J. Kent, Adam K. Wheatley, Sharon R. Lewin, Kanta Subbarao, Amy Chung, Marc Pellegrini, Trent Munro, Terry Nolan, Steven Rockman, David C. Jackson, Damian F.J. Purcell, Dale I. Godfrey

**Author notes:** These authors contributed equally (Georgia Deliyannis, Nicholas A. Gherardin). These authors contributed equally (David C. Jackson, Damian F.J. Purcell).

## Abstract

The SARS-CoV-2 global pandemic has fuelled the generation of vaccines at an unprecedented pace and scale. However, many challenges remain, including: the emergence of vaccine-resistant mutant viruses, vaccine stability during storage and transport, waning vaccine-induced immunity, and concerns about infrequent adverse events associated with existing vaccines. Here, we report on a protein subunit vaccine comprising the receptor-binding domain (RBD) of the ancestral SARS-CoV-2 spike protein, dimerised with an immunoglobulin IgG1 Fc domain. These were tested in conjunction with three different adjuvants: a TLR2 agonist R4-Pam2Cys, an NKT cell agonist glycolipid α-Galactosylceramide, or MF59® squalene oil-in-water adjuvant. Each formulation drove strong neutralising antibody (nAb) responses and provided durable and highly protective immunity against lower and upper airway infection in mouse models of COVID-19. We have also developed an RBD-human IgG1 Fc vaccine with an RBD sequence of the highly immuno-evasive beta variant (N501Y, E484K, K417N). This ‘beta variant’ RBD vaccine, combined with MF59® adjuvant, induced strong protection in mice against the beta strain as well as the ancestral strain. Furthermore, when used as a third dose booster vaccine following priming with whole spike vaccine, anti-sera from beta-RBD-Fc immunised mice increased titres of nAb against other variants including alpha, delta, delta+, gamma, lambda, mu, and omicron BA.1 and BA.2. These results demonstrated that an RBD-Fc protein subunit/MF59® adjuvanted vaccine can induce high levels of broad nAbs, including when used as a booster following prior immunisation of mice with whole ancestral-strain Spike vaccines. This vaccine platform offers a potential approach to augment some of the currently approved vaccines in the face of emerging variants of concern, and it has now entered a phase I clinical trial.

## Introduction

Widespread vaccination across all the world’s regions is viewed as our best chance to overcome, or at least live comfortably with, COVID-19 infection. There are several vaccines now approved for use in many countries, mostly based on the induction of neutralising antibodies (nAb) to spike (S) protein using the ancestral SARS-CoV-2 S sequence. These include mRNA vaccines (eg. BNT162b2^1^ and mRNA-1273^2^), viral vector vaccines (eg. ChadOx1^3^, Ad26.COV2.S^4^, whole inactivated virus vaccines (eg. BBIBP-CorV^5^ and CoronaVac^6^) and protein subunit vaccines (NVX-CoV2373^7^, ZF2001^8^). These vaccines have demonstrated efficacy ranging from ∼50% to 95% against symptomatic infection with the ancestral SARS-CoV-2 strain (reviewed in ^9,10^). However, SARS-CoV-2 continues to evolve, leading to the repeated emergence of variants of concern (VOC), that can reduce vaccine efficacy. This is highlighted by the most recent VOC, omicron and its subvariants, that are highly infectious and immuno-evasive even in populations that have received up to four doses of existing vaccines ^11-14^.

It is likely that the world will require ongoing boosters to minimise viral transmission, infection and severe disease, and to limit emergence of new SARS-CoV-2 variants. This has prompted the question of whether immunity to SARS-CoV-2 can be improved through the development of vaccines tailored to VOC, with several vaccine candidates in trials ^15-20^. To achieve a more comprehensive global coverage and limit the spread and/or development of VOC, current licensed vaccines require improvement. Improvements in vaccine design regarding storage and transportability are required to ensure that vaccines can be distributed through regions that lack the infrastructure for cold-chain storage and transport. As well as overcoming potential problems that may confound the efficacy of vaccines that are modified against SARS-CoV-2 VOC, including rare but serious adverse reactions, including thrombosis with thrombocytopenia syndrome (TTS) associated with some viral vector vaccines (reviewed in ^21^) and myocarditis and pericarditis associated with some mRNA vaccines (reviewed in ^22^). Yet, perhaps the greatest obstacle to re-directing the immune responses to VOC is the phenomenon of immunological imprinting, also known as original antigenic sin, a well-recognised problem in the context of influenza infection and vaccination (reviewed in ^23^). Emerging evidence suggests that this same problem limits the boosting effect of whole S-based booster vaccines that incorporate VOC S protein sequences, presumably because the immune system prefers to target common epitopes shared between the mutant spike vaccine and the ancestral strain to which all approved vaccines are aligned ^24,25^ (reviewed in ^26^). Interestingly, some studies have suggested that exposure to the beta VOC, either by infection or vaccination, provides broader neutralising antibodies against a range of VOC including omicron ^19,20,27,28^.

In an effort to circumvent some of these problems, we developed a vaccine candidate based on the SARS-CoV-2 receptor-binding domain (RBD), generated as an Fc fusion protein, to facilitate multimeric presentation to the immune system while also engaging Fc receptor (FcR)+ antigen-presenting cells for enhanced immunological priming. While an RBD vaccine does not necessarily overcome the imprinting problem, there will be no distraction of the immune system to Spike epitopes outside of the RBD, which are highly likely to generate non-neutralising antibody responses, whereas RBD epitopes, whether VOC-specific or shared with the ancestral strain, are more likely to induce neutralising antibodies ^29^. This RBD vaccine induced high titres of RBD-specific antibodies, including high nAb titres, in mice following a prime and boost regimen. Immunity induced by this vaccine was durable and provided complete protection against heterologous strain virus challenge in mouse models of SARS-CoV-2 infection. Furthermore, this vaccine is highly adaptable to incorporate RBD sequence changes, and we have engineered versions corresponding to several VOC RBDs. For clinical testing, we engineered a beta variant version of our RBD human IgG1-Fc vaccine, which based on recent studies, may yield broader immune responses to VOCs including omicron ^19,20,27,28^. Indeed, this beta RBD vaccine induced a potent nAb response against the beta SARS-CoV-2 virus, and protected mice against infection with this strain, as well as a mouse-tropic version of the ancestral SARS-CoV-2 strain, and generated strong nAb against other VOC. To replicate the current community situation where most individuals have been vaccinated with spike-based vaccines, we also tested the ability of our beta-RBD-Fc vaccine as a heterologous boost in mice that have previously received two doses of SARS-CoV-2 S protein vaccine. These results showed that our beta RBD vaccine as a third injection provided a stronger immune boost for generating RBD-targeting antibodies, including nAb, compared to a third ancestral or beta variant whole spike vaccine boost. We also provide evidence that this boosted immunity was also increased for other VOC including the ancestral strain, alpha, gamma, delta, kappa and omicron BA.1 and BA.2. A phase I clinical trial of our vaccine in individuals previously primed and boosted with licenced SARS-CoV-2 vaccines is currently underway.

## Results

### Strong immunogenicity and protection in mice using a SARS-CoV-2 RBD-Fc vaccine

The SARS-CoV-2 receptor-binding domain (RBD) is responsible for viral binding to the host-cell receptor, Angiotensin-converting enzyme-2 (ACE-2) and facilitating virus entry^30^. This small region of the virus is the target of over 90% of nAb following SARS-CoV-2 infection ^29,31,32^. For these reasons, we engineered a soluble form of the SARS-CoV-2 RBD as an RBD-Fc dimer fusion protein, fusing the N334-P527 of the RBD to the core hinge region of mouse IgG1 via a short serine/glycine linker (Figure 1A and Supplementary Figure 1), to use as a protein subunit vaccine against SARS-CoV-2 virus. We confirmed the activity of this protein by demonstrating that it specifically bound to ACE2-transduced HEK-293T cell lines but not control HEK-293T cells transduced with an irrelevant protein (Supplementary Figure 1).

**Figure 1.**
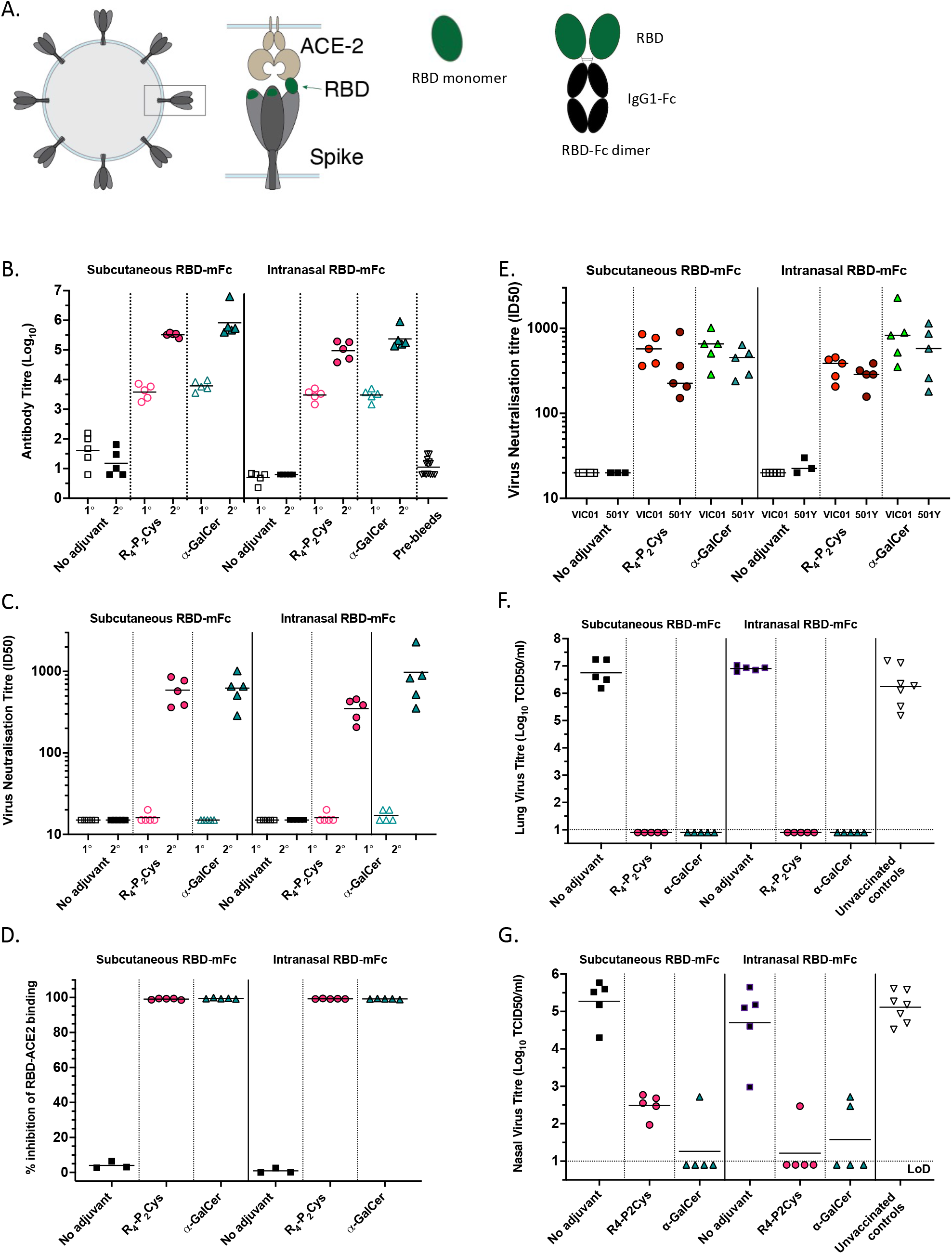
Immunogenicity and protection in mice using a SARS-CoV-2 RBD-Fc vaccine. **(A)** Cartoon diagram of SARS-CoV-2 virus covered in spike proteins at its surface and highlighting the receptor-binding domain (RBD) which binds ACE2 to elicit viral entry into cells. Cartoon diagram of RBD monomer and RBD-Fc dimer proteins. **(B - G)** BALB/c mice (n = 5/group) were vaccinated subcutaneously or intranasally with 10µg ancestral RBD-mFc only (No adjuvant), or 10µg RBD-mFc with R4-Pam2Cys (0.3 nmole) or α-GalCer (0.2µg) on day 0 and 29. **(B)** Total WT RBD antibody titres in primary (1°, day 13) and secondary (2°, day 41) sera measured by ELISA. **(C)** Neutralisation of ancestral index strain VIC01 via micro-neutralisation assay. Neutralisation titres of primary (1°, day 27) and secondary (2°, day 41) sera. **(D)** Percent inhibition of RBD binding to ACE2 using sVNT assay on secondary (day 41) sera from mice. **(E)** Comparison of neutralisation of index strain VIC01 to VIC2089 (N501Y, D614G) via micro-neutralisation assay using secondary (day 41) sera. Note - VIC01 datapoints are repeated from 1C secondary sera samples. **(F and G)** Titre of virus in the lungs and nasal turbinates of mice challenged with VIC2089 (N501Y, D614G). Mice were aerosol challenged with VIC2089 SARS-CoV-2 strain, 74 days after the second immunisation. Age and sex matched unvaccinated control BALB/c mice were also challenged at the same time. Three days later, challenged mice were killed, and the titre of infectious virus in the lungs **(F)** and nasal turbinates **(G)** of individual mice was determined by titrating homogenised lung and nasal supernatants on Vero cell monolayers and measuring viral cytopathic effect (CPE) 5 days later. LoD: Limit of detection. Horizontal lines depict means.

Groups of BALB/c mice were immunised with this vaccine via subcutaneous or intranasal routes, either in the absence of adjuvant, or with two experimental adjuvants: R4-Pam2Cys, a TLR-2 agonist^33^, or α-galactosylceramide (α-GalCer), an NKT cell agonist^34^. Both adjuvants are known to promote dendritic cell maturation and enhanced adaptive immunity to protein antigens. Both subcutaneous and intranasal administration of two doses of the RBD-Fc dimer vaccine induced high anti-RBD antibody (Figure 1B) and nAb titres, as measured by an *in vitro* SARS-CoV-2 microneutralisation assay (Figure 1C) and by a surrogate virus neutralising test (sVNT)^35^ (Figure 1D), but only in the presence of adjuvant, with little distinction between the two adjuvants tested. Using a microbead based assay, we determined that sera from the immunised mice were capable of binding to a broad range of RBDs with point mutations (Supplementary Figure 2A) including the N501Y mutation that represents the alpha variant RBD. Furthermore, anti-sera from the immunised mice neutralised a naturally arising D614G/N501Y SARS-CoV-2 clinical variant (VIC2089) that we had isolated and adapted for use in a microneutralisation assay (Figure 1E). This variant is capable of productive infection in mice (Supplementary Figure 2B)^36^, allowing us to develop a challenge model to test the protective capabilities of the vaccine. Mice immunised via either the subcutaneous or intranasal routes, samples from which are depicted in Figures 1B-E, were challenged with VIC2089 and lungs and nasal turbinates harvested 3 days post-challenge. Complete protection against lung infection was observed in mice immunised with the RBD-Fc vaccine in combination with either R4-Pam2Cys or α-GalCer adjuvant, whereas no protection was observed with protein alone (no adjuvant group) (Figure 1F). There was also a clear reduction in viral titres in the upper airways (nasal turbinates), from 10^5^ TCID^50^/ml down to less than 10^3^ TCID50/ml, in RBD-Fc + adjuvant-immunised mice, with more than half of the mice showing no detectable virus (Figure 1G). In the subcutaneous immunisation group, α-GalCer cleared the virus from nasal turbinates in 4/5 mice whereas R4-Pam2Cys markedly reduced, but did not clear, the virus following this administration route. These data demonstrate that the RBD-Fc dimeric vaccine provides strong protection against SARS-CoV-2 infection in mice following a prime/boost regimen, but only when used in the presence of adjuvant, and both R4-Pam2Cys and α-GalCer adjuvants were highly effective at inhibiting infection in this site, particularly following intranasal administration.

### Intramuscular injection of the RBD vaccine provides strong and durable immunity in mice

As most vaccines used in humans are given via the intramuscular route, we next tested the efficacy of the RBD-Fc vaccine injected into the quadricep of mice, in the presence of R4-Pam2Cys adjuvant. Furthermore, to ensure that the vaccine was immunogenic in more than one mouse strain, we compared this response in both C57BL/6 and BALB/c mice. This route of injection resulted in high total RBD-specific (Figure 2A) and nAb titres, as measured by microneutralisation assay (Figure 2B). When these mice were challenged with SARS-CoV-2 virus, we were unable to detect virus in lungs of either strain (Figure 2C). Virus was also undetectable in nasal turbinates of all C57BL/6 mice and two of the BALB/c mice, but three of the BALB/c mice still had some virus, albeit lower than any of the unimmunised BALB/c mice (Figure 2D). It is noteworthy that the protection was durable, as mice were challenged 75 days after the boost. We also observed that the RBD-Fc vaccine with R4-Pam2Cys was highly immunogenic in both mouse strains via the subcutaneous route, however, C57BL/6 mice showed a weaker response compared to BALB/c mice when immunised via the intranasal route (data not shown). Further testing via the intranasal route is warranted but has not been performed. In summary, these data suggest that this RBD vaccine, introduced via the intramuscular route, reduces viral burden in both lower and upper airways, at least in mice, and if this also applies to humans, this may also help to reduce SARS-CoV-2 transmission.

**Figure 2.**
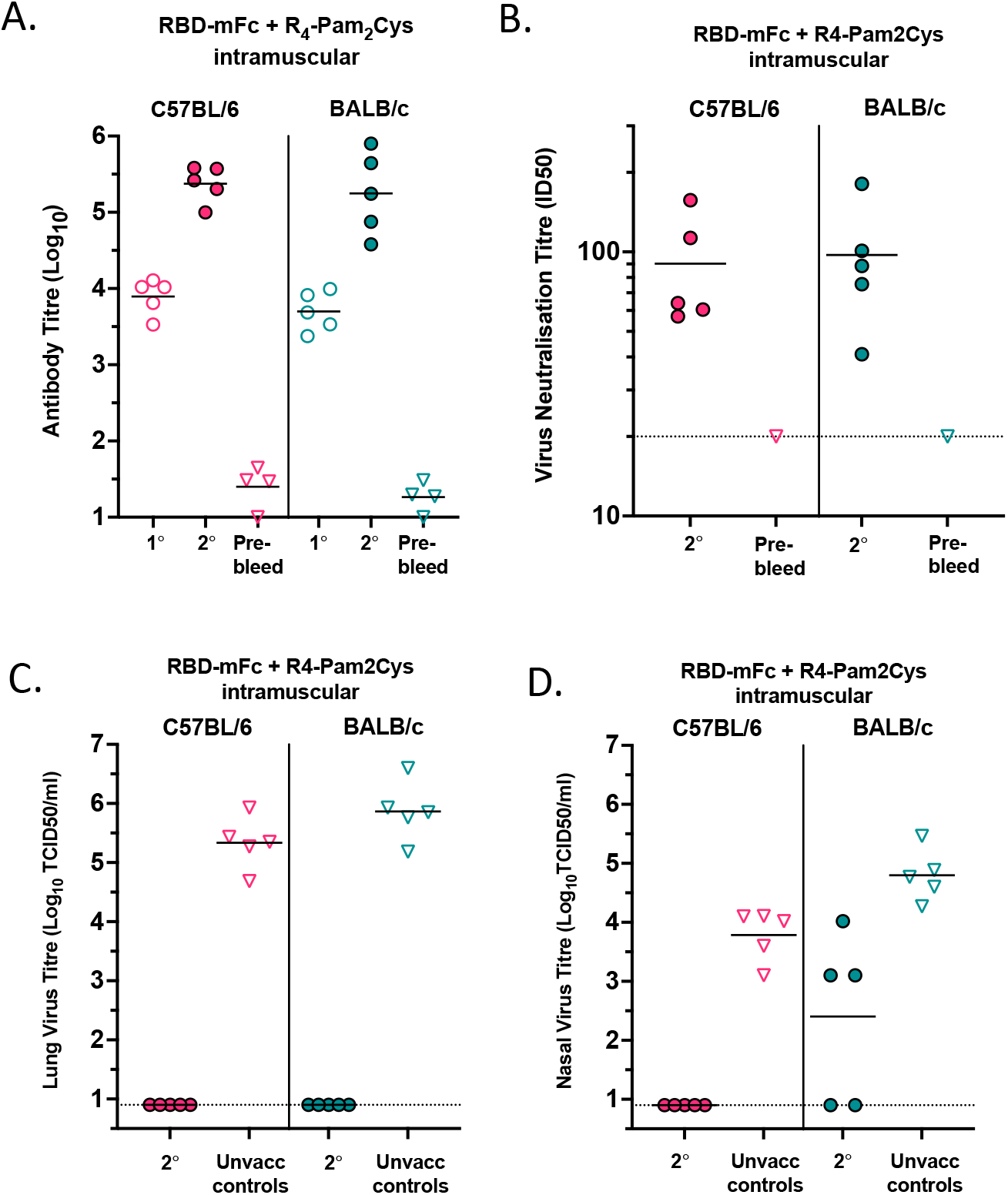
Immunogenicity and protection in BALB/c and C57BL/6 mice inoculated intramuscularly with a SARS-CV-2 RBD-Fc vaccine. **(A)** Total anti-RBD antibody titres in primary (1°, day 13) and secondary (2°, day 27) sera from mice inoculated via the intramuscular route with 10µg WT RBD-mFc with 0.3 nmoles of R4-Pam2Cys on days 0 and 13. **(B)** Neutralisation of WT Index strain VIC01 via micro-neutralisation assay. Neutralisation titres of secondary (2°, day 27) sera were determined. **(C and D)** Mice were aerosol challenged with VIC2089 on day 89 (75 days after the second immunisation). Age and sex matched unvaccinated control C57BL/6 and BALB/c mice were challenged at the same time. Three days after challenge, mice were killed, and the titre of infectious virus in the lungs **(C)** and nasal turbinates **(D)** of individual mice were determined. Horizontal lines depict means.

### RBD-Fc dimeric vaccine provides superior immunity to RBD monomeric vaccine

Given that other RBD protein-based vaccines, including RBD monomers ^37^ and single chain dimers ^8,38^ have used a third dose in mice to generate a strong neutralising immune response, we assessed whether the Fc-dimer form of our vaccine was responsible for the high level of immunity observed with just two doses. We directly compared our chimeric RBD-mouse IgG1 Fc dimeric protein to a simple RBD monomeric protein domain (Figure 3A), immunising mice with these vaccines combined with R4-Pam2Cys adjuvant in a prime-boost regimen. Both forms of RBD vaccine were injected via the intramuscular route, and total anti-RBD antibody tested at d28 after the primary injection and d13 after the secondary. nAb were tested 28 days following the secondary (Figure 3B-C). The results demonstrated that the RBD-Fc dimer form of our vaccine provided superior total and nAb responses following both the prime and boost stages. We then tested the potential of these two versions of RBD vaccines to protect in the SARS-CoV-2 VIC2089 (N501Y) mouse challenge model. While both forms of the RBD vaccine (+ adjuvant) were capable of suppressing virus infection in lungs, 2/10 mice immunised with RBD monomer still showed high viral loads in lungs (Figure 3D), and all these mice had high viral titres in nasal turbinates (Figure 3E). In contrast, mice immunised with the RBD-Fc vaccine (+ adjuvant) showed complete protection against infection in both lungs and nasal turbinates, with no detectable virus in either sample for 10/10 mice (Figure 3D and E). These data suggest that the RBD-Fc dimer vaccine will be superior to an RBD monomer vaccine, particularly when trying to prime naïve B cell responses against novel epitopes such as those present in SARS-CoV-2 VOC.

**Figure 3.**
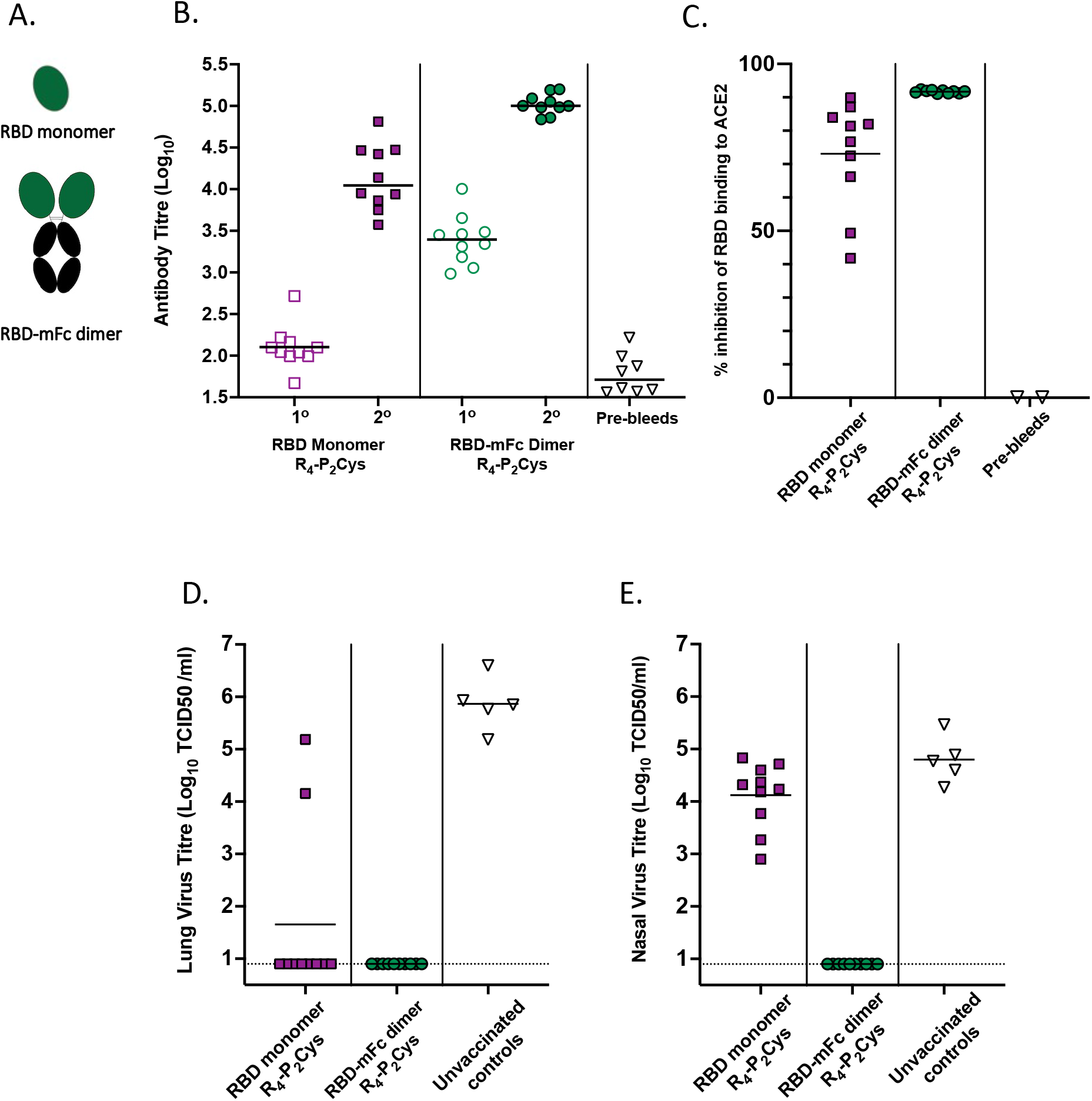
Immunogenicity and protective efficacy of RBD monomer and RBD-Fc dimer vaccines. **(A)** Cartoon diagram of RBD monomer and RBD-Fc dimer proteins. **(B)** Total WT RBD antibody titres in primary (1°, day 28) and secondary (2°, day 41) sera from BALB/c mice inoculated via the intramuscular route with 10µg RBD monomer or 10µg RBD-mFc dimer in the presence of 0.3 nmoles R4-Pam2Cys. **(C)** Percent inhibition of RBD binding to ACE2 using sVNT assay on secondary (day 41) sera. **(D and E)** Titre of virus in the lungs and nasal turbinates of mice challenged with VIC2089. Mice were aerosol challenged 20 days after the second immunisation. Age and sex matched unvaccinated control BALB/c mice were also challenged at the same time. Unvaccinated control mouse data is repeated from Figure 2C and D because the virus challenge experiments for vaccinated mice in figures 2 and 3 were conducted at the same time using the same group of control mice. Three days after challenge, mice were killed, and the titre of infectious virus in the lungs **(D)** and nasal turbinates **(E)** of individual mice was determined. Horizontal lines depict means.

### Adaptation of the RBD-Fc protein vaccine for clinical use

As the goal was to progress our vaccine to human clinical trials, we generated an RBD-human Fc dimer protein (using the human IgG1 Fc region) and compared the immunogenicity of this protein to the RBD-mouse Fc dimer in both BALB/c and C57BL/6 strains. Mice were immunised with a prime boost regimen, via the intramuscular route, with RBD-mouse or human Fc vaccines combined with R4-Pam2Cys. As shown in Figure 4A, the human Fc form of the RBD vaccine induced a similar level of anti-RBD antibodies to the mouse Fc form, albeit slightly lower in C57BL/6 mice. In both mouse strains, the mouse and human Fc form of the RBD vaccine induced durable antibody responses that were generally maintained at a constant level up to day 115 (87 days post-boost) (Figure 4A).

**Figure 4.**
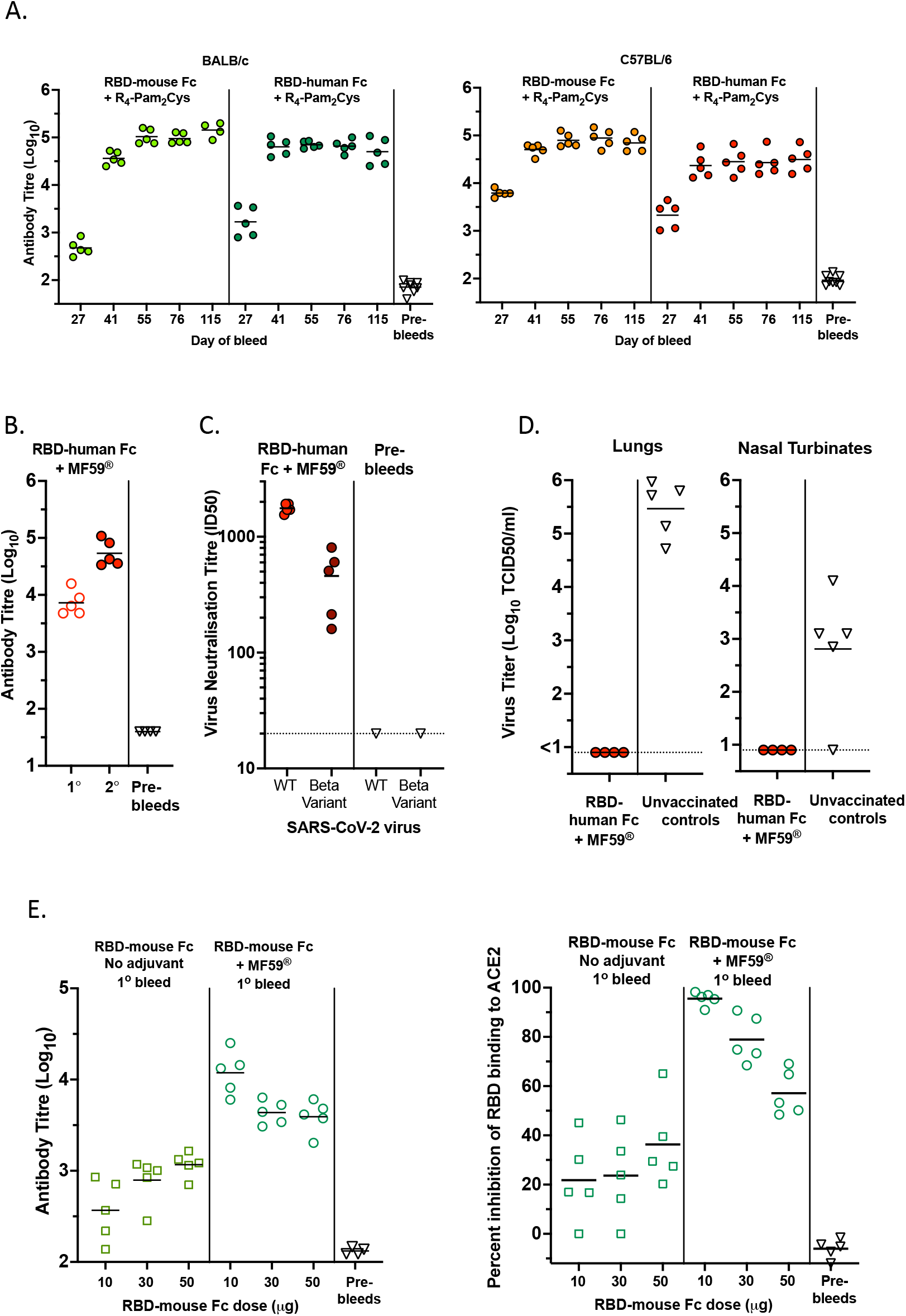
Immunogenicity and protective efficacy of RBD-human Fc vaccine. **(A)** Total anti-RBD antibody titres in primary (day 27) and secondary (day 41, 55, 76 and 115) sera from BALB/c and C57BL/6 mice vaccinated intramuscularly on day 0 and 28 with 10µg RBD-mouse Fc or 10µg RBD-human Fc, combined with 0.3 nmoles of R4-Pam2Cys. **(B and C)** Total and nAb responses in C57BL/6 mice vaccinated intramuscularly with 10µg RBD-human Fc + MF59® on day 0 and 21. **(B)** Total anti-RBD antibody titres in primary (1°, day 20) and secondary (2°, day 33) sera. **(C)** Neutralisation of ancestral (WT) index strain VIC01, or Beta variant B.1.351, via micro-neutralization assay. Neutralisation titres of secondary (day 45) sera were determined. **(D)** The C57BL/6 mice vaccinated intramuscularly on days 0 and 21 with 10µg WT RBD-hFc formulated with MF59® were aerosol challenged with VIC2089 60 days after the second immunisation. Age and sex matched unvaccinated control C57BL/6 mice were also challenged at the same time. Three days after challenge, mice were killed, and the titre of infectious virus in the lungs and nasal turbinates of individual mice was determined. **(E)** Immunogenicity of higher antigen doses of RBD-mouse Fc formulated with MF59® after a single vaccination. C57BL/6 mice were primed via the intramuscular route with 10, 30 or 50µg of RBD-mFc in the absence (No adjuvant) or presence of MF59®. Sera collected on day 43 post-priming were assayed for total anti-RBD antibody levels measured by ELISA, and neutralising activity assessed as the percent Inhibition of RBD binding to ACE2 using the sVNT. Horizontal lines depict means.

We also investigated whether a human-IgG1 Fc form of our vaccine would work with a clinically approved and widely used oil-in-water adjuvant, MF59®. The RBD-human-Fc dimer was mixed with MF59® adjuvant and mice immunised via the intramuscular route with a prime-boost regimen. This vaccine formulation resulted in high levels of total RBD-specific antibody (Figure 4B) and nAb including against the immuno-evasive beta VOC (Figure 4C) and provided complete protection in mice challenged with the VIC2089 strain, with no detectable virus in lungs or nasal turbinates (Figure 4D).

To determine whether scaling up the dose in mice might further enhance the antibody response, the RBD-mFc dimer was tested at a range of doses (10, 30 and 50µg per injection), with or without MF59® adjuvant, as a single injection. Whereas increasing doses made little difference to the low responses observed in the absence of adjuvant, surprisingly, an inverse relationship was detected between the dose of RBD-mFc vaccine with MF59® and the amount of total and nAb (Figure 4E). This suggests that higher doses may not be advantageous and may even result in suboptimal immune responses. Furthermore, a single 10µg dose of this vaccine with MF59® adjuvant was able to induce a strong nAb response.

### A SARS-CoV-2 beta variant RBD-human IgG1Fc vaccine

SARS-CoV-2 VOC such as the beta, delta and omicron strains can at least partially resist vaccine-induced immunity mediated by ancestral strain-based vaccines^11-14,16^. In preparing vaccine for a phase I clinical trial, we reproduced the RBD-hFc using a Chinese hamster ovary (CHO) cell line approved for production of clinical grade recombinant proteins. We took this opportunity to generate a version of the protein that carried three mutations: N501Y, E484K and K417N, corresponding with the highly immuno-evasive beta variant which was prevalent at the time when we commenced vaccine production of the clinical-grade material in early 2021.

As before, mice were immunised via the intramuscular route with the beta RBD-human Fc vaccine, alongside other mice immunised with the ancestral (wildtype (WT)) RBD-human-Fc vaccine described above. Considering the inverse dose relationship observed in Figure 4E, we also tested a lower dose range including 10, 3, 1 and 0.3 µg doses. Antibody titres were measured against both WT and beta RBDs antigen targets in ELISA and we observed similar antibody titres from mice that had received either vaccine (Figure 5A and B). Also unexpectedly, while the 10µg dose produced the highest average antibody titres, the lowest 0.3µg dose was still very immunogenic with titres of approximately 10^4^. As a more direct measure of the immunogenicity and protective potential of the beta RBD vaccine, we directly compared neutralisation of the WT and the beta variant strains of SARS-CoV-2 by microneutralisation assay. These results demonstrated that the mean neutralising titres of serum samples from mice immunised with the beta variant RBD vaccine were moderately higher against the beta variant virus than against the ancestral virus (Figure 5C).

**Figure 5:**
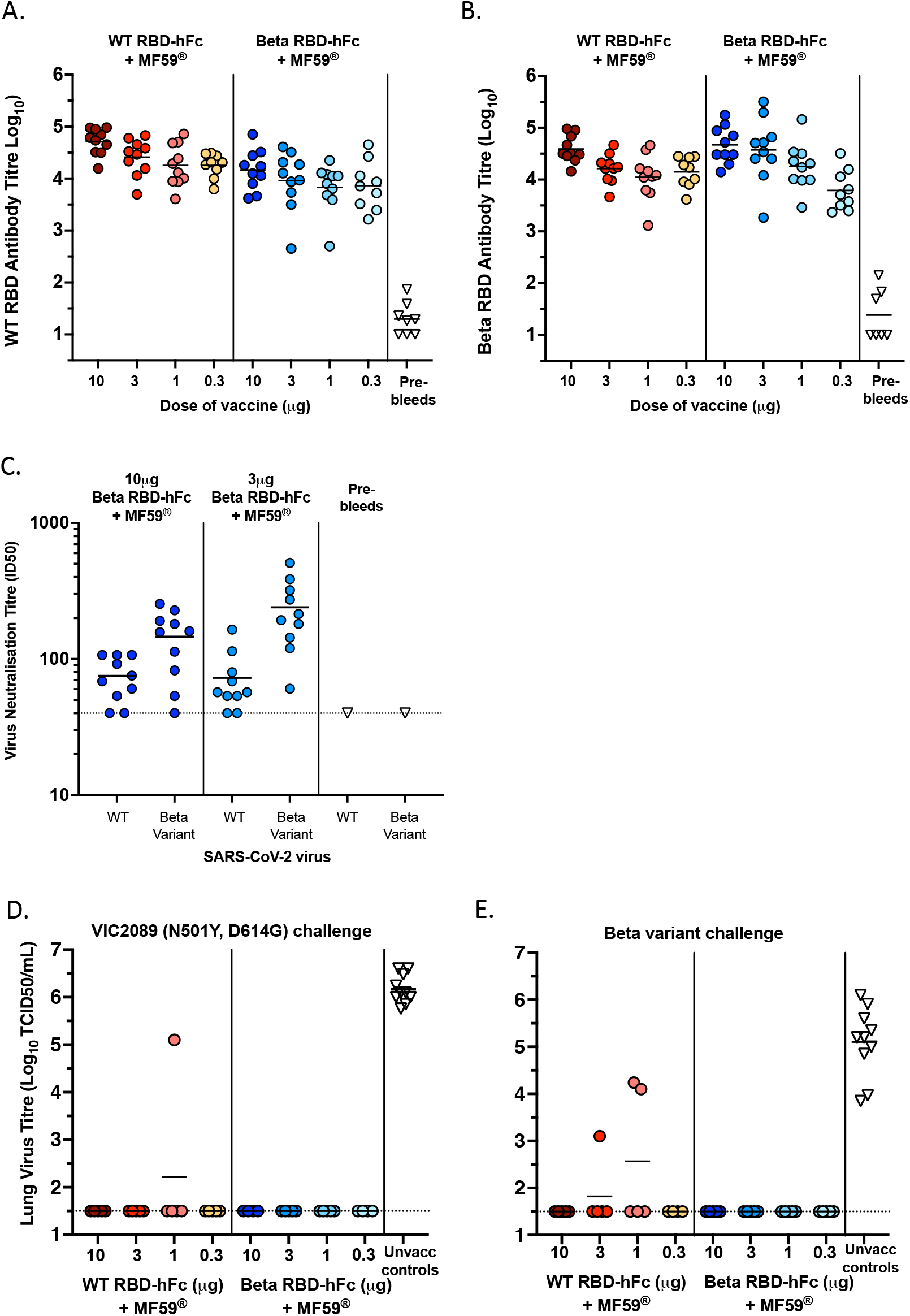
Immunogenicity and protective efficacy of a SARS-CoV-2 beta variant RBD-human Fc vaccine. Ancestral (WT) RBD-specific antibody titres **(A)** and beta RBD-specific antibody titres **(B)** in secondary sera (day 56) from C57BL/6 mice vaccinated intramuscularly on days 0 and 21 with (10, 3, 1 or 0.3µg) of WT RBD-hFc or Beta RBD-hFc in the presence of MF59®. **(C)** Neutralisation of Index strain VIC01 (WT) or Beta variant B.1.351 via micro-neutralisation assay. Neutralisation titres of secondary (day 33) sera were determined for serum samples from mice immunised with 10 and 3 µg of beta-RBD-hFc. **(D-E)** Mice were aerosol challenged with VIC2089 **(D)**, or Beta variant B.1.351 **(E)** 55 or 62 days respectively, after the second immunisation. Age and sex matched unvaccinated control C57BL/6 mice were also challenged on each of the corresponding days. Three days after challenge mice were killed, and the titre of infectious virus in the lungs of individual mice were determined. Horizontal lines depict means.

Protective efficacy against lung infection was assessed in the mouse SARS-CoV-2 challenge model using VIC2089 (N501Y) and the beta VOC (B.1.351) as challenge strains. On day 76 of the study (55 days after the boost) 5 of the 10 mice from each vaccination group immunised with 10, 3, 1 or 0.3µg of either WT RBD-hFc or beta RBD-hFc in the presence of MF59® were challenged with VIC2089 (Figure 5D). On day 83 of the study (62 days after the boost) the remaining 5 mice in each group were challenged with the beta variant B.1.351 (Figure 5E).

Complete protection against lower airway infection after challenge with either VIC2089 or beta variant was indicated in all C57BL/6 mice vaccinated intramuscularly with 10, 3, 1 or 0.3µg of beta RBD-hFc in the presence of MF59®. Of the mice vaccinated with WT RBD-hFc + MF59®, 19 of the 20 mice challenged with VIC2089 and 17 of the 20 mice challenged with beta variant had no detectable virus in their lungs (Figure 5D-E).

### Rat and hamster studies

The immunogenicity of the beta RBD-hFc vaccine with or without MF59® adjuvant in rats was also assessed as an alternative model to mice. As part of a pre-clinical toxicology study, Sprague Dawley Rats were vaccinated intramuscularly on days 0, 22 and 43 with saline, 50μg of beta RBD-hFc or 50μg of beta RBD-hFc with MF59® adjuvant. Blood samples were collected within 2 days (day 44-45), or 13 days (day 56) after thevaried levels of resistance to third vaccination and assessed for antibody responses against both ancestral (WT) and beta variant RBDs by ELISA (Figure 6A). The beta RBD-hFc vaccine with MF59® was highly immunogenic and, with the exception of only one rat, the other 29 rats produced high antibody titres against both RBD antigens. In contrast, only 3 of the 30 rats that were immunised with beta RBD hFc without MF59® produced detectable antibodies against WT and beta RBDs. These samples were also analysed using the sVNT assay for nAb, clearly showing that all rats that received beta RBD-hFc vaccine with MF59® produced nAb, with all but one sample still neutralised out to a 1:80 dilution (Figure 6B).

**Figure 6:**
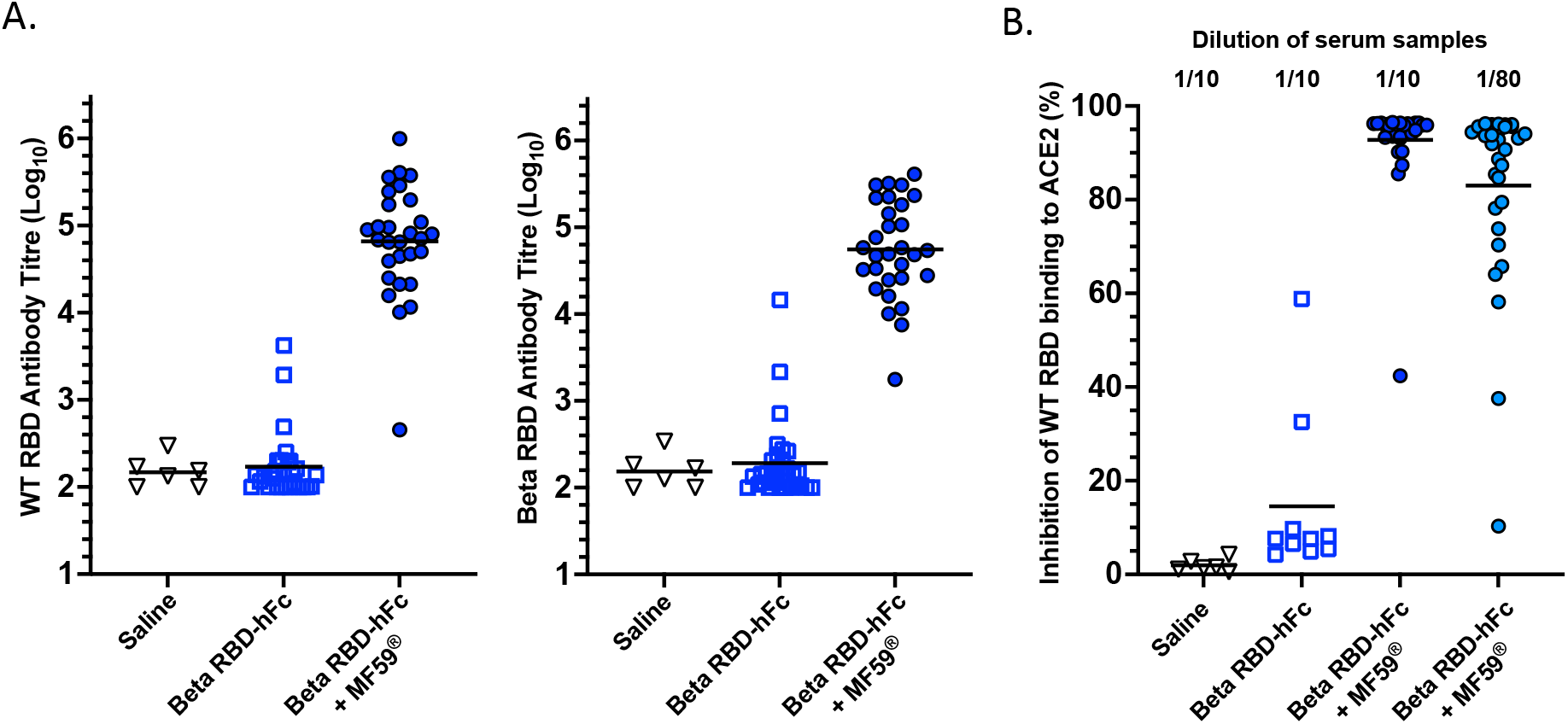
Immunogenicity of beta RBD-hFc in rats. Rats were vaccinated intramuscularly on days 0, 22 and 43 with saline, 50μg of beta RBD-hFc or 50μg of beta RBD-hFc + MF59® adjuvant. Blood samples were collected within 2 days (day 44-45), or 13 days (day 56) after the third vaccination and assessed for WT SARS-CoV-2 RBD-specific and beta SARS-CoV-2 RBD-specific antibody responses by ELISA **(A)** and nAb measured by sVNT assay at 1/10 and 1/80 dilutions **(B)**. Horizontal lines depict means.

We also tested our RBD-human Fc vaccines in hamsters. In the first experiment, performed at The University of Hong Kong, we tested the ancestral RBD-hFc vaccine with either MF59® or R4-Pam2Cys adjuvant. While both formulations induced an antibody response in hamsters (Supplementary Fig 3A), it was not as strong as the responses in mice, so a third dose was administered. While most hamsters produced anti-RBD antibody (Supplementary Fig 3A), for unknown reasons, there were also positive readings from PBS-injected hamsters following the third vaccination. Neutralising antibodies were detected in most immunised hamsters, but not in the PBS-injected hamsters. However, three hamsters from the 30µg RBD-hFc + MF59® group and two from the 10µg RBD-hFc + MF59® group had low or undetectable nAb titres (Supplementary Fig 3B). Of these, one hamster in the 30µg group, and all in the 10µg group showed detectable nAb titres 4 days post SARS-CoV-2 virus challenge whereas the PBS-injected group remained negative for nAb. All hamsters that received RBD-hFc with R4-Pam2Cys produced nAb. Three weeks after the third vaccination, hamsters were challenged with SARS-CoV-2 virus and killed 4 days later to measure viral load in lung and nasal tissues (Supplementary Fig 3C). Four of five hamsters in the group that received 10µg RBD-hFc with MF59® demonstrated clear protection with no detectable or low lung or nasal virus titres. Curiously, of hamsters that received the higher 30µg dose of RBD-hFc with MF59®, three exhibited low or negligible viral titres (<1000 PFU/ml), but two showed no reduction in viral load. This is reminiscent of the data from above (Figure 4E) where a higher dose of the RBD-hFc with MF59® vaccine may be inferior to a lower 10µg dose of vaccine. Of the hamsters that received RBD-hFc with R4-Pam2Cys at 30 and 10µg, all had low or undetectable viral titres in lung and nasal tissues.

A second hamster study was conducted at Colorado State University, USA, where it was possible to challenge hamsters with both a WT strain (WA-01/USA) and beta variant (B.1.351) of SARS-CoV-2. In this experiment, hamsters were vaccinated with 30 or 10µg of the WT RBD-hFc + MF59® or 30, 10 or 3µg beta RBD-hFc + MF59®, receiving 3 intramuscular doses on days 0, 21 and 63. Fourteen to 21 days after each vaccination, antibody titres were measured against both WT and beta variant RBDs as coating antigens in ELISA (Supplementary Fig 4A). Antibody responses were detectable in most hamsters after the first immunisation and were clearly increased after the second and third immunisation, although the antibody titres were not as high as we observed for mice and rats. We observed similar antibody responses against the two RBD antigens in hamsters vaccinated with beta RBD-hFc with MF59®, however hamsters vaccinated with WT RBD-hFc with MF59® displayed slightly higher average responses against the WT RBD antigen. Neutralising Ab titres against WT SARS-CoV-2 virus were assessed 2 weeks after the third vaccine dose using the plaque reduction neutralisation test (PRNT). Results demonstrate that 7 of 10 hamsters immunised with WT RBD-hFc dimer, and 15 of 30 hamsters immunised with beta RBD-hFc dimer, had detectable nAb responses (Supplementary Fig 4B). While there was a high degree of variability between hamsters, the 30µg vaccine dose appeared to be no better than the 10µg dose at driving total Ab or nAb responses. Hamsters were then challenged with either WT or beta variant SARS-CoV-2 virus, oropharyngeal swabs collected every day for 3 days and hamsters euthanised after 3 days for lung virus titre assessment. Titres of virus in oropharyngeal swabs, and viral load/mg of cranial and caudal lung tissue and turbinate tissue, on day 3 post-challenge, is presented. Consistent with the moderate and variable nAb response observed (Supplementary Fig 4B), the immunised hamsters also varied levels of resistance to viral challenge (Supplementary Fig 4C and D). For the WT virus challenge, the mean viral load in the oropharyngeal swabs of control hamsters administered saline declined rapidly (by two orders of magnitude) between day 1 and day 3, with one having undetectable virus at day 3, which left little room for improved responses with the vaccinated hamsters. Nonetheless, in day 2 swabs, there was more than 10-fold lower mean viral loads in hamsters immunised with 30µg and 10µg WT RBD-hFc (282 and 212 PFU/swab) compared to saline controls (5510 PFU/swab). By day 3, three hamsters treated with 30µg WT RBD-hFc had undetectable, and two had low, virus titres (mean 13 PFU/swab) while the 10µg WT RBD-hFc group all had low but detectable virus (mean 42 PFU/swab). For hamsters immunised with 10 or 3µg beta RBD-hFc and challenged with WT virus, by day 2 they also had mean reductions of more than 10-fold (530 and 176 PFU/swab respectively compared to 5510 PFU/swab in controls). In contrast, the mean viral titre in the 30µg group was not much lower than the control group (3263 versus 5510). By day 3, mean virus titres were low in all groups (29, 18 and 61 PFU/swab, respectively) with several at or below the lower limit of detection, but as mentioned above, the saline control treated hamsters also had low titres (77 PFU/swab) by day 3 (Supplementary Fig 4C).

Hamsters challenged with the beta variant (B.1.351) virus sustained high viral loads in the PBS-treated group over the 3 days post-challenge (day 3 mean 1200 PFU/swab) and vaccine-induced protection was apparent in day 3 swab samples. Thus, all hamsters immunised with beta RBD-hFc, except for one in the 3µg group, had lower viral loads, below the range of the control group (mean PFU/swab of 110, 32 and 250 from the 30, 10 and 3µg treated groups, respectively) (Supplementary Fig 3E). Results from day 3 cranial and caudal lung tissue gave similar evidence, with modest to strong reductions in pfu in some but not all groups (Supplementary Fig 3E). The nasal turbinate data only showed a strong (>10-fold) reduction in viral titres at the 10µg dose. Collectively, these hamster data show partial protection in a dose-dependent manner, and interestingly, they suggested that the 10µg dose was more effective than the 30µg dose for both the WT and beta variant vaccines in the hamster models.

Thus, while the rat and hamster studies both demonstrated immunogenicity and induction of protective nAb, the results from hamsters were more variable compared to the rat and mouse models. A possible limitation of these hamster studies is that, as shown previously, RBD-based vaccines may be less immunogenic in hamsters, for reasons that are unclear ^39^. This limitation does not apply to humans where RBD-based vaccines are effective ^8,40,41^.

### Heterologous boosting with RBD-Fc vaccines

Given nAb titres wane within months with currently approved vaccines, particularly against SARS-CoV-2 variants that can at least partially evade pre-existing immunity, booster vaccines are now being used to enhance immunity to levels that exceed those after the second dose ^17,42-46^. Some of these boosters are heterologous (different from the priming vaccine) and some are designed to target VOC although in most cases, these do not appear to markedly improve the immune response to those VOC when compared to boosting with the ancestral strain-based vaccine ^15-20^.

We investigated whether the beta RBD-hFc vaccine could act as a heterologous boost in animals that had previously been primed, or primed and boosted, with whole WT S protein vaccine plus MF59® adjuvant. Groups of mice were primed with ancestral (WT) S vaccine, then boosted with various combinations of WT S vaccine, beta S vaccine, WT RBD-hFc vaccine or beta RBD-hFc vaccine, each in the presence of MF59® adjuvant. For the first part of this experiment, we examined the response in mice, measuring anti-WT RBD-reactive antibodies after priming (day 21) and boosting (day 35) (Figure 7A). Strong primary responses were observed in the sera of all groups after priming, although in this experiment the beta RBD-hFc primed mice had moderately lower mean titre relative to the other groups. Markedly increased levels of ancestral RBD-specific antibodies were detected in sera collected 2 weeks after the second dose (day 35) in all groups of mice, regardless of whether they were boosted with WT S, beta S, WT RBD-Fc or beta RBD-Fc vaccines (Figure 7A). Post-boost serum samples from these mice were then tested against either the WT RBD, or beta or delta RBDs, in a separate ELISA (Figure 7B). All groups showed very strong and comparable responses against each variant RBD. This demonstrates that the RBD-Fc vaccines provide at least comparable boosting ability to the whole S vaccines. In this experiment, we separated data from the 4 groups of 5 mice that were primed and boosted with WT S vaccine (labelled 1-4, Figure 7A) as they were subsequently used for a heterologous third dose boost experiment (below).

**Figure 7:**
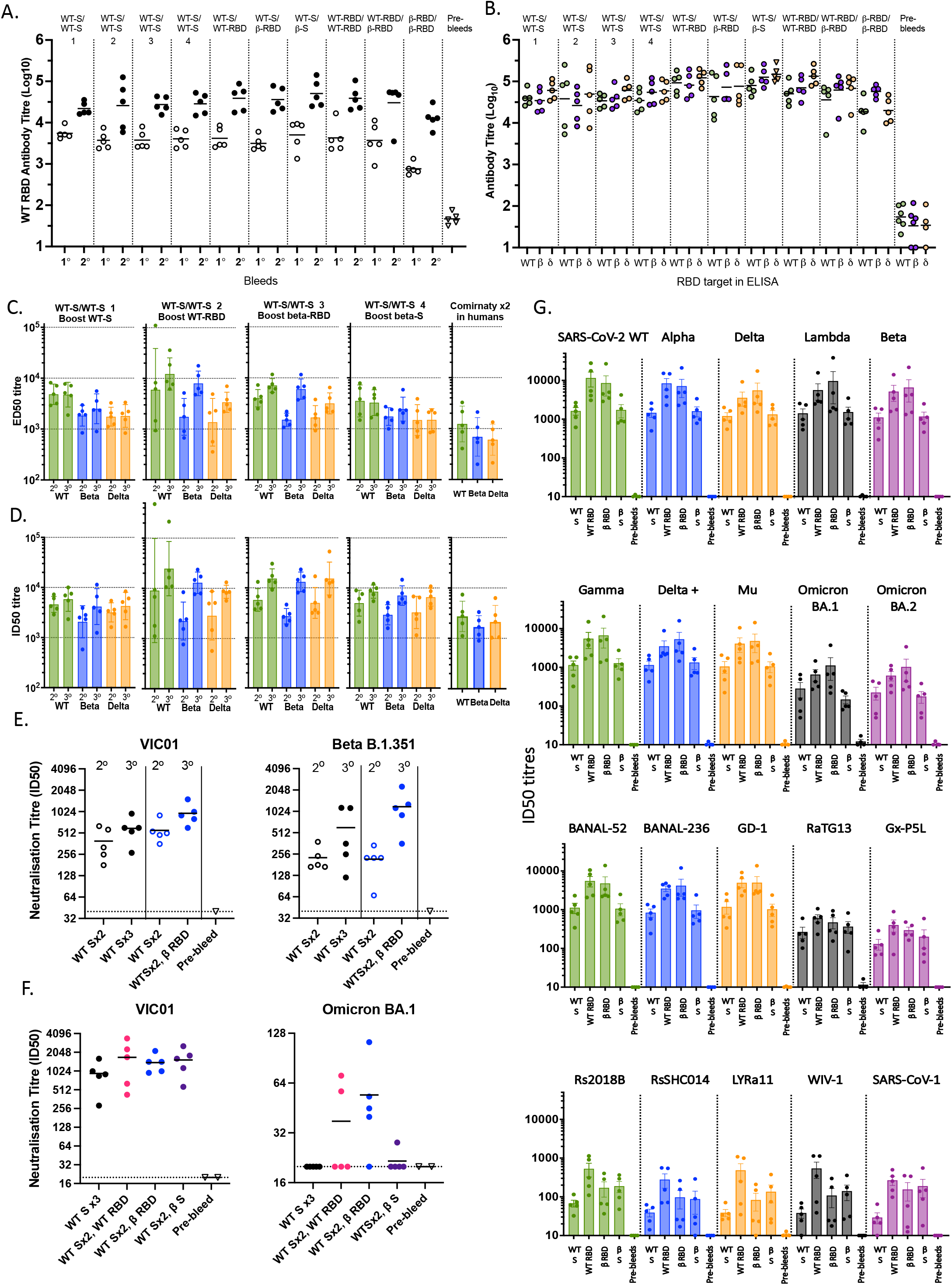
Heterologous boosting with RBD-Fc vaccines. **(A-B)** C57BL/6 mice (n=5) were vaccinated intramuscularly on day 0 and 21 with either the spike antigens or the RBD-hFc antigens as indicated above each column. More than half of the mass of RBD-Fc vaccine is Fc, therefore mice were boosted with 10µg of RBD-Fc vaccine (∼4.5µg of virus protein) or 4.5µg S vaccine. All antigens were administered with MF59®. **(A)** Total WT RBD-specific antibody titres determined by ELISA, in primary (1°, day 21) and secondary (2°, day 35) sera. **(B)** Total antibody titres to WT RBD, beta variant (β) RBD, and delta variant (δ RBD in secondary (day 56) sera determined by ELISA. **(C-G)** The 4 groups of C57BL/6 mice vaccinated intramuscularly on day 0 and 21 with 4.5µg of the WT-spike with MF59® were boosted on day 70 with either 10µg WT RBD-hFc, 10µg beta RBD-hFc, 4.5µg of WT-spike or 4.5µg of beta-spike in the presence of MF59®. **(C)** Multiplex bead-based assay was used to assess IgG antibody responses to WT, beta and delta RBD monomers in secondary (2°) sera collected 35 days following the second immunisation and tertiary sera (3°) collected 16 days following the third immunisation. Half-maximal effective dilution (ED50) is indicated for each serum sample and **(D)** in an RBD-ACE2 multiplex inhibition assay for their ability to inhibit the binding of ACE2 to WT-RBD, beta variant (β) RBD and delta variant (δ RBD where half-maximal Inhibitory dilution (ID50) is indicated for each serum sample. **(E)** A selection of these serum samples were further assessed in a microneutralisation assay for nAb responses against WT VIC01 or beta variant B.1.351. **(F)** A selection of serum samples taken from a later timepoint (day 30) following the third immunisation were tested in an omicron-BA.1 variant microneutralisation assay using TMPRSS2 overexpressing Vero cells, alongside a VIC01 microneutralisation assay using conventional Vero cells. **(G)** Samples from day 30 following the third immunisation were tested in a multiplex sVNT inhibition assay for their ability to inhibit the binding of ACE2 to 20 different CoV RBD variants, including SARS-CoV-2 VOCs (Alpha, Beta, Gamma, Delta, Delta+, Lambda, Mu, Omicron BA.1 and Omicron BA.2), animal SARS-CoV-2-related coronaviruses (BANAL-52, BANAL-236, GD-1, RaTG13 and GX-P5L), animal SARS-CoV-1 related coronaviruses (Rs2018B, LYRa11, WIV-1, RsSHC014) and SARS-CoV-1. Half-maximal Inhibitory dilution (ID50) is indicated for each serum sample. Bars depict means and SEM (A, B, E, F, G) or geometric mean and geometric SD (C and D).

Because many people have already been primed and boosted with whole S vaccines, we next tested the ability of the RBD-hFc vaccines to provide a second boost (third injection) using the 4 groups of 5 mice that had been primed and boosted with ancestral S vaccine (from the experiment presented above in Figure 7A and B). These mice were boosted on day 70 (49 days after their second dose) with either WT S protein, WT RBD-hFc, beta RBD-hFc, or beta S protein, each with MF59® adjuvant. Secondary boost sera collected 5 weeks (day 56) following two doses of the WT S protein (2° bleed) and tertiary sera collected 16 days (day 86) following the third immunisation (3° bleed) were assessed using a multiplex bead-based assay for binding to WT RBD, beta RBD and delta RBD (Figure 7C). These data suggested that the third injection with RBD vaccine, either WT or beta, provided a stronger boost with higher mean ED50 titres compared to a third injection with whole WT or beta S vaccines. It is noteworthy that for all vaccinated mouse samples in this experiment, regardless of which boost they received, the mean titres were considerably higher than antibody titres from 5 adult human donors previously primed and boosted with the BNT162b2 (*Comirnaty*^*TM*^) vaccine and tested in the same assay (Figure 7C). We also examined nAb responses from these mice using a multiplex bead-based RBD-ACE2 binding inhibition assay (Figure 7D). Strong neutralising activity against the WT RBD, beta RBD and delta RBD (indicated as half-maximal inhibitory dilution (ID50)) was observed in secondary sera from all groups of mice when compared to those from samples from the *Comirnaty*^*TM*^ vaccinee human donors. Two weeks after the third vaccination, mean nAb levels were highest in mice that had received a third dose of an RBD-hFc vaccine relative to the mice that were boosted with spike-based vaccines (Figure 7D) although there was not much difference between the WT and the beta RBD vaccine boosts. An expanded version of these data, incorporating both binding ED50 titres and neutralising ID50 titres for other mutant RBDs including S477N, E484K, L452R, alpha, gamma and kappa sequences is provided in Supplementary Figure 5A and B. In general, the RBD vaccine boosts induced at least a moderately higher mean nAb titre than the whole S vaccine boost. Furthermore, these mean ED50 and ID50 readings were all higher those from the *Comirnaty*^*TM*^ vaccinee human donors.

A selection of samples from the mice described above were assessed for nAb against both WT (VIC01) and beta VOC (B.1.351) SARS-CoV-2 viruses using a SARS-CoV-2 virus micro-neutralisation assay. Four sets of mouse samples were tested: 1. primed and boosted with WT S + MF59®; 2. The same mice from 1. that were again boosted on day 70 with WT S + MF59®; 3. primed and boosted with WT Spike + MF59®; 4. The same mice from 3. that were again boosted on day 70 with beta RBD-hFc + MF59® (Figure 7E). These results demonstrated that the beta RBD-hFc + MF59® boost induced higher mean nAb responses against both the WT virus and the beta variant strain compared to pre-boost levels. By this assay, the boost provided by the beta RBD-hFc + MF59® was at least equal to, if not better than, the boost provided by the third dose of WT spike + MF59® vaccine, for both WT and beta viruses.

We next tested serum samples from these mice for their ability to neutralise the omicron VOCs (using a later bleed, 30 days after the third dose boost). In this assay, we tested mice that had received third dose boosts with WT S, WT RBD-hFc, beta RBD-hFc and beta S. We initially performed a micro-neutralisation assay where omicron BA.1 virus was added at a standard dose of 100 TCID50/well, however, in this assay, all samples from mice immunised with 3 doses of ancestral S showed no neutralising activity above the lower limit of detection (Figure 7F). Similarly, all but one sample from beta S-boosted mice had undetectable nAb against BA.1. In contrast, 2/5 samples from WT RBD-boosted mice and 4/5 from beta RBD-boosted mice had clearly detectable nAb titres. As the dynamic range of this assay appeared to be too low to compare nAb titres between the four groups, we also tested samples in a similar assay, set up with a lower amount of BA.1 virus (10 TCID50/well) for improved sensitivity (Supplementary Figure 6). While the trends were similar to the previous assay (Figure 7F), this assay revealed a mean titre of 112 for the WT-S 3-dose group; and 51 for the beta S vaccine-boosted group, versus 203 for WT-RBD-Fc vaccine-boosted mice and the highest mean nAb titre of 290 for beta RBD-boosted mice.

As new VOC are constantly emerging, including omicron sub-lineages BA.2, BA.3, BA.4 and BA.5 and now sub-sub lineages such as BA.4.6 and BA.5.9, it is increasingly clear that we require a vaccine that can provide broad immunity against SARS-CoV-2 variants, ideally including future variants. Therefore, we tested serum samples from the heterologous third dose boost experiments described above, in a different RBD-ACE2 binding inhibition (sVNT) bead assay carrying a 20-plex panel of CoV-derived RBDs including some of the SARS-CoV-2 VOC RBDs described above, as well as Omicron BA.1 and BA.2, plus a panel of bat and pangolin CoVs and SARS-CoV-1 RBD^47^ (Figure 7G). Samples from 30 days after the third dose boost were tested in this assay which should have allowed more time for affinity maturation of RBD-reactive B cells. These data highlighted the breadth of nAb induced by the ancestral and beta RBD-hFc vaccines. Thus, mean ID50 readings were between 3 and 7-fold higher for samples from the WT and beta RBD boosted mice for all the SARS-CoV-2 RBDs tested, including ancestral (WT), alpha, beta, gamma, delta, delta+, lambda, mu, omicron BA.1, omicron BA.2 (Figure 7G). Similarly, the RBD boosts resulted in increased mean titres against bat CoVs BANAL-52, BANAL-236 and the pangolin CoV GD-1. Compared to the WT S boost, there were also increased mean titres for the WT RBD boosted mice against bat CoVs RS2018B, RsSHCO14, LyRa11, WIV-1, and pangolin CoV GxP-5L (Figure 7G). Interestingly, for RaTg13, Gx-P5L, Rs2018B, RsSHCO14, LYRa11, WIV-1 and SARS-CoV-1 RBDs, mean nAb titres in serum samples from beta RBD-boosted mice were similar to beta S-boosted mice (Figure 7G).

Collectively, these data suggest that the RBD-targeted vaccine can work well as a heterologous boost following priming and boosting with whole S-vased vaccine. Moreover, these data suggest that the RBD-targeted vaccines can augment nAb responses toward RBD epitopes that are shared between the S vaccine prime/boost and the RBD vaccine third dose boost. Many of these epitopes are common to all VOC, including omicron, which also carries the mutations N501Y and K417N that are shared between omicron and beta RBDs. In contrast, the whole beta S vaccine boost may be more likely to target many of the shared non-RBD epitopes that are less likely to be neutralising. Taken together, these studies suggest that the beta-variant RBD-human IgG1-Fc + MF59® vaccine is a good candidate for heterologous boosting of subjects previously primed or primed and boosted with whole spike vaccines. This vaccine is currently being assessed in a phase I clinical trial (ClinicalTrials.gov Identifier: NCT05272605).

## Discussion

The production of vaccines, via a variety of platforms, seek to protect against SARS-CoV-2 infection. The current vaccine program has had a major impact on the severity of COVID-19 in communities with access to these vaccines. However, in light of waning immunity and the continuing emergence of new VOC with increasing degrees of resistance to vaccine or prior infection-induced immune responses, vaccine boosters are now widely used. While these are capable of boosting immunity, the breadth of protection and durability of booster responses against new variants still appears to be limited, particularly in the face of the omicron variants^11-14,18^. To address the problems posed by emerging VOC, variant-targeted or bivalent vaccine boosters are being trialled, but at best, these only appear to provide roughly 2-fold immune boost over the ancestral strain-targeting vaccines^17-20^. This is potentially due, at least in part, to the phenomenon of immunological imprinting, or original antigenic sin^14,23,24,48^ (reviewed in ^26,46^) which restricts immune responses against spike-based vaccines to epitopes that are shared with the ancestral strain. It is therefore prudent to explore new vaccine platforms that provide potential ways around these problems, whilst still offering the potential for rapid adaptability and repeated immunisation without cumulative side effects. A major advantage of an RBD vaccine is that it focusses the immune response on the most important target of the virus in terms of nAb neutralisation, because even though the RBD only comprises roughly 15% of the S protein, 90% of more nAb are directed toward this region of the virus ^29,31,32^. It follows that if a variant-targeted booster version of the RBD vaccine is introduced, there will be far fewer original ancestral strain epitopes, most of which are non-neutralising, in the booster RBD vaccine, compared to a variant whole S booster vaccine. While imprinting may still occur with an RBD variant vaccine, at least the antigens that are common between the old and new strains will all be directed to the RBD region and therefore more likely to promote nAb, regardless of whether they are targeting ancestral or VOC RBD epitopes.

Our evidence using the beta RBD-Fc vaccine described in this paper is noteworthy in this regard, because we demonstrated that the response when this is used as a third dose boost, following priming and boosting with an ancestral (WT) strain S vaccine, results in higher mean antibody levels, including nAb, compared to a third dose boost with either WT, or a beta variant, S vaccine. While this was apparent for the beta VOC to which our RBD vaccine is targeted, encouragingly it was also observed for other variants, including alpha, gamma, delta, delta+, lambda, mu, omicron BA.1, omicron BA.2 and some bat and pangolin CoVs. While delta shares none of the RBD mutations with beta, omicron shares two (N501Y and K417N), plus in both beta and omicron, E484 is mutated, however, in beta it is mutated to E484K, while in omicron it is mutated to E484A. Therefore, we suggest that the beta RBD-Fc vaccine provided a strong boost response mainly because it is amplifying and focussing existing S vaccine driven immunity to the RBD region of the virus, and this may further benefit from alignment of some of the RBD mutations with the VOC being targeted.

Based on the mouse data, the RBD-hFc vaccine appears to be most effective at low µg doses (1-10µg), and this combined with high production yields of ∼1g per litre (not shown), means that if a similar low dose range is optimal in humans, this candidate vaccine platform could be a very efficient means for high level production. The Fc dimeric form of this vaccine produced in mammalian cells means that production and purification can utilise industry manufacturing platforms already established for rapid and large-scale production and purification of antibody-based therapeutics, avoiding delays associated with complex production of other vaccine platforms. Furthermore, stability studies thus far have shown that this vaccine is stable for up to at least 12 months at 2-8°C, and at least 2 weeks at 37°C (not shown), it will be highly amenable to transportation and storage in regions where a reliable frozen/cold chain infrastructure is lacking.

Several RBD protein vaccines are in various stages of development and testing(https://covid19.trackvaccines.org/vaccines)^8,37,40,41,49-58^, and one, ZF2001^8,50^, is now approved for emergency use in several countries, including China, Uzbekistan, Indonesia and Columbia (https://covid19.trackvaccines.org/vaccines/27/). This is a dimeric RBD vaccine where two RBD subunits are linked via an engineered single-chain construct and administered with Alum adjuvant as a three-dose schedule^8^. A similar dimeric RBD vaccine, combined with Alum plus Neisseria meningitidis outer membrane vesicles, is also in phase I clinical trials^51^. Another RBD-human IgG1-Fc dimer, fused to IFN-α and an MHC class-II binding element, combined with Alum adjuvant, has also recently been through phase I and II clinical trials^40,52^ and beta and delta VOC versions of this are in preclinical testing^53^. This vaccine was well tolerated and induced nAb in all subjects, generally exceeding those of convalescent serum. Interim results from a phase I/II trial of an RBD-human IgG1 Fc vaccine (ancestral strain) with montenide oil-in-water adjuvant in a two-dose prime-boost schedule provided evidence of strong neutralising antibody responses with low levels of reactogenicity^41^. Another approach used to develop an RBD-based vaccine was to fuse two RBDs to a hepatitis B surface antigen (PreS) plus Alum, which generated robust responses in rabbits and in a single human volunteer^54^. Others have developed RBD monomeric vaccines with Alum or Alum-based adjuvants also administered as three-dose schedules^37,55^ one of which is in human clinical trials^55^. A recent pre-clinical study of an “RBD mosaic” nanoparticle vaccine including RBDs from 8 different sarbecoviruses, adjuvanted with *Addavax*^*TM*^ (an MF59®-like adjuvant), demonstrated impressively broad immunity against SARS-CoV-2 VOCs as well as other Sarbecoviruses including SARS-CoV-1^56^. One study described a monomeric RBD vaccine that, when used with *AddaVax*^*TM*^, proved to be a potent vaccine when used in a heterologous prime/boost approach in animals, where whole spike protein vaccine with *AddaVax*^*TM*^ was used to either prime the response followed by the RBD boost, or vice versa^57^. Taken together, these studies demonstrate that RBD-based protein vaccines are immunogenic in humans. This supports our view that our uniquely formulated beta-variant RBD-Fc dimer/MF59® adjuvanted vaccine will work well as a heterologous booster subsequent to S-based vaccination in humans, particularly considering the increased breadth of immune responses that appear to be promoted by the beta variant^19,20,27,28^.

Our evidence suggests that the Fc dimer design of our vaccine is a key to why our vaccine can promote strong immune responses after only two injections in naïve mice, with a 10-fold increase in RBD-specific Ab titres in mice given 2 doses of RBD Fc dimer compared to mice given RBD monomer. There may be several reasons for why the Fc-dimer works better than the monomer: The Fc-dimeric construct is likely to bind to FcR+ antigen-presenting cells such as dendritic cells, macrophages and B cells, presenting a multimeric array of RBD protein in the context of professional antigen presenting cells, to enhance RBD-specific B cell activation. Furthermore, the ligation of FcR may activate the FcR-expressing cells, enhancing immunity to the vaccine^59^. Regardless of the reason, the superior ability of an Fc-conjugated vaccine to prime new immune responses may be crucial for enhancing responses using a VOC-targeted boosters, because novel epitopes are generated in VOC that require priming of new VOC-specific immune cells. In addition to the FcR binding, we tested our vaccine with three different adjuvants including R4-Pam2Cys, α-GalCer and MF59® and determined that each adjuvant augmented the immune response to our vaccine. MF59® was selected for further investigation because it is a widely used oil-in-water adjuvant that has been injected into more than a hundred million people in association with influenza vaccines^60^. This adjuvant has a proven safety profile with little if any reactogenicity reported, aside from transient local pain at the injection site. We have shown that MF59® works exceptionally well as an adjuvant in association with this RBD protein vaccine, and it seems likely that it will be at least as effective in humans. Moreover, as the human IgG1 Fc based vaccine appeared to be very potent in mice, we would expect that this format should also enhance RBD immunity in humans because the human IgG1 Fc region will interact strongly with human FcR+ cells, further enhancing the immune response to this RBD-Fc vaccine.

There are several limitations to this study. While the vaccine has been extensively tested in mice, including mouse and human Fc versions, as well as rats and hamsters, we await the results of clinical trials to determine how immunogenic our vaccine is in humans. Based on results from phase I/II clinical trials of other experimental ancestral strain RBD-Fc protein vaccines used in prime-boost regimen in humans, albeit with different adjuvants ^40,41,52^, and our preclinical data, we anticipate that our vaccine used as a fourth dose boost will augment nAb responses in humans. While hamsters also responded to a prime-boost regimen with our RBD vaccines, the response was more varied and not as robust as was observed in mice and rats, which may reflect findings in a recent report of limitations in the extent to which hamsters respond to RBD-based vaccines ^39^. Another potential limitation is that, compared to whole spike vaccines, RBD vaccines which obviously contain only some of the T cell epitopes present throughout S protein, may not induce as broad T cell immune responses as whole S vaccines. While we did not measure T cell responses in this study, this region of the S protein is known to contain immunodominant T cell epitopes^61-63^ and it is clear that this vaccine is capable of driving isotype-switching leading to high levels of IgG production. Importantly, while the vaccine is capable of priming and boosting in an immunologically naïve setting, given that many people have either been immunised or infected or both, we are primarily interested its potential to augment nAb responses as a booster vaccine. As increased nAb responses is a correlate of protection against VOC viruses ^64^, our mouse-based data suggests that this will be a strength of our vaccine. Lastly, our lead vaccine candidate is based on the RBD from the beta VOC. While this is no longer a circulating VOC, having since been replaced by others and most recently omicron VOCs, our findings are that the beta RBD-hFc boost drives a broad nAb response that spans all of the SARS-CoV-2 VOCs we tested. Furthermore, studies with human samples have shown that exposure to the beta VOC was superior to the ancestral strain at driving broadly cross-reactive nAb, including those that can target omicron^19,20,27,28^.

In summary, we describe an RBD-Fc protein subunit vaccine that induces potent nAb and complete and durable protection against lower and upper airway infection in a mouse model of SARS-CoV-2 infection. Furthermore, we show that an RBD-Fc vaccine incorporating three mutated residues from the beta VOC can promote potent nAb responses that target the beta variant virus in *in vitro* microneutralisation assays, while still driving a broad response that can also neutralise the ancestral strain in vitro and in a mouse challenge model. And importantly, the breadth of this response suggests that this vaccine can also boost nAb against other VOC including delta, and omicron BA.1 and BA.2 as well as other variants. This vaccine is now in phase I clinical trial as a fourth dose boost. The future of COVID-19 vaccines, including those specific for VOC, will require rapid and scalable manufacturing. The RBD-Fc fusion protein should be a suitable candidate for commercialisation. The manufacturing of this product using mammalian cells and primary capture using Protein-A-based chromatography allows utilisation of existing commercial biotechnology infrastructure used for products like recombinant monoclonal antibodies. These facilities could rapidly output large numbers of doses, including for the developing world, given the promising stability profile without the need for frozen storage and transport.

## Materials and Methods

### Recombinant RBD Protein constructs

#### RBD-mouse IgG1 Fc-fusion proteins (WT RBD-mFc)

Recombinant DNA fragments encoding the truncated RBD of the WT isolate of SARS-CoV-2 (N334-P527; genbank accession NC_045512) were synthesised between a 5’ AgeI and a 3’ BamHI cloning site (GeneArt Gene Strings, Thermo Scientific). These were then cloned into the mammalian expression vector pHLSec, fusing the C-terminus of the RBD to the Fc-domain of mouse IgG1 from the core hinge region through to the C-terminal lysine via a GSGSG linker.

#### RBD-human IgG1 Fc-fusion protein (WT RBD-hFc)

Recombinant DNA fragments encoding the truncated RBD as above, fused via a GSGSG linker to the Fc domain of human IgG1 from the core-hinge region to the C-terminal lysine followed were codon-optimised for mammalian expression (GeneArt Gene Strings, Thermo Scientific), synthesised between a 5’ NheI and a 3’ XhoI cloning site (IDT), and cloned into the mammalian expression vector pHLSec.

#### Beta variant RBD-human IgG1 Fc-fusion (Beta RBD-hFc)

For clinical-grade product, the WT construct above was subcloned into pXC-17.4 for production of stable CHO cells, after HindII and EcoRI cloning sites were introduced via PCR. PCR mutagenesis was then used to introduce K417N, E484K and N501Y, correlating with the RBD mutations of the beta VOC.

#### RBD monomers

Recombinant DNA fragments encoding the truncated WT RBD as above or Delta isolate (https://www.cdc.gov/coronavirus/2019-ncov/variants/variant-info.html) were codon-optimised for mammalian expression (GeneArt Gene Strings, Thermo Scientific), synthesised between a 5’ NheI and a 3’ BamHI cloning site (IDT) and cloned into the mammalian expression vector pHLSec to incorporate a C-terminal 6-HIS tag. The beta RBD monomer (residues 332-532) used for ELISA assays was a gift from Professor Heidi Drummer, and Dr Rob Center, Burnet Institute, Melbourne, Australia.

### Protein Expressions

All RBD proteins, with the exception of the beta RBD human IgG1 Fc fusion protein and beta RBD monomer, were expressed by transient transfection of Expi293F cells using ExpiFectamine 293 Transfection Kits as per manufacturer’s instructions (ThermoFisher Scientific). Proteins were harvested on day six. RBD-Fc proteins were purified from supernatants by Protein A Sepharose (CL-4B, Cytiva) and RBD monomers were purified by Ni-NTA resin (HisPur, Thermo Scientific). All proteins were further purified by gel filtration size exclusion chromatography: RBD-Fc using a Superdex-200 column (Cytiva), and RBD monomers using a Superdex S75 column (Cytiva). All proteins were sterile filtered and stored at -80°C prior to use.

For RBD-beta human IgG1 Fc fusion protein, stable cell lines were generated using the Lonza GS Xceed® System (Lonza, Basel, Switzerland). CHOK1SV GS-KO® cells were transfected via electroporation with linearized GS expression vector encoding Beta-RBD-Fc as per manufacturer’s instructions (GS Xceed® manual, Version 06 2019). Enriched minipools were selected using 50 µM L-Methionine sulfoximine (MSX) over a period of 3-4 weeks. Lonza’s abridged fed-batch shake flask screen was performed to assess the minipools and choose the lead pool.

For beta RBD monomer, proteins were expressed in Expi-293F cells. Four days after transfection, tissue culture supernatants were clarified and target proteins were purified by IMAC using Talon metal affinity resin (Clontech Laboratories) following the manufacturer’s recommendations. The eluted proteins were subject to gel filtration using a Superdex 200 16/600 column (GE Healthcare) with PBS as the liquid phase. Fractions corresponding to monomeric RBD were pooled and concentrated in Amicon Ultra 30kDa devices (Merck) prior to use.

### Experimental mice

Male C57BL/6 and BALB/c mice aged 6-10 weeks were used in this study. Mice were maintained in the Biological Research Facility in the Department of Microbiology and Immunology at the University of Melbourne. All animal experimentation was conducted in accordance with institutional regulations following review and approval by the University of Melbourne Animal Ethics Committee.

### Human samples

Human plasma samples were obtained from individuals two-weeks following second dose with BNT162b2 vaccine. One plasma sample was also collected from a convalescent breakthrough infected individual after their third BNT162b2 vaccine dose. The study protocols were approved by the University of Melbourne Human Research Ethics Committee (2021-21198-15398-3, 2056689), and all associated procedures were carried out in accordance with approved guidelines. All participants provided written informed consent in accordance with the Declaration of Helsinki.

### Vaccination of mice

C57BL/6 and BALB/c mice were vaccinated either via the subcutaneous route at the base of the tail (100μl), intranasally to the upper respiratory tract (15μl) or intramuscularly into the right caudal thigh musculature (50μl). For all vaccination routes, mice were anaesthetised by isoflurane inhalation using an anaesthetic machine with a controlled oxygen flow rate, and a controlled anaesthetic vapour flow concentration. RBD protein antigens were either administered in the absence of adjuvant or administered in the presence of 0.3 nmoles of R4-Pam2Cys, 0.2µg α-GalCer or (50% vol/vol) MF59® adjuvant. Mice were primed on day 0 and boosted on either day 14, 21 or 28 (as indicated in figure legends). Submandibular venous bleeds of mice were carried prior to the first vaccination (pre-bleed), just prior to the second injection (1° bleed), and at multiple timepoints following the second injection (2°bleeds).

### Enzyme-linked immunosorbent assay (ELISA) for measurement of RBD-specific antibody responses

WT and variant RBD-specific total antibody responses in the sera of mice pre- and post-inoculation were investigated by ELISA using the RBD monomer from either the WT, Beta or Delta variant strain. Flat bottom 96 well maxisorp plates (ThermoFisher Scientific) were coated with 50μl/well of RBD monomer at a concentration of 2 μg/ml in Dulbecco’s phosphate buffered saline (DPBS; Gibco Life Technologies). Plates were incubated overnight at 4°C in a humidified atmosphere. Unbound antibody was removed, and wells were blocked with 100 μl/well of 1% bovine serum albumin (BSA fraction V, Invitrogen Corporation, Gibco) in PBS for 1-2 hours before washing 3 times with PBS containing 0.05% v/v Tween-20 (PBST). Serial dilutions of mouse sera were added to wells and left to incubate overnight at room temperature. After washing, bound Ab was detected using horseradish peroxidase (HRP)-conjugated rabbit anti-mouse Ig Abs (Dako, Denmark). The detection antibody was incubated for 1 hour at room temperature in a humidified atmosphere and the plates then washed five times with PBST. 100μl of tetramethylbenzidine substrate (TMB, BD Biosciences) was then added to each well and the reaction was stopped after 5-7 minutes by the addition of 100 μl/well of 1M orthophosphoric acid (BDH Chemicals, Australia). A Labsystems Multiskan microplate reader (Labsystems, Finland) was used to measure the optical density (OD) of each well at wavelengths of 450 nm and 540 nm. The titers of Ab are expressed as the reciprocal of the highest dilution of serum required to achieve an OD of 0.3 which represents at least five times the background level of binding.

### In vitro microneutralisation assay (mNT)

An in vitro micro-neutralisation assay measured the level of SARS-CoV-2-specific nAb in sera of immunised or infected mice. SARS-CoV-2 isolates used in the microneutralisation assay were propagated in Vero cell cultures and stored at -80°C. Flat-bottom 96-well plates were seeded with Vero cells at 2 × 10^4^ cells/well the day before assay. Serial 2-fold dilutions of heat-inactivated sera were incubated with 100 TCID50 (50% tissue culture infectious dose) of SARS-CoV-2 for 45 min and residual virus infectivity was assessed in quadruplicate wells of Vero cells. Plates were incubated at 37°C and viral CPE was read on day 5. The dilution of serum that completely prevented CPE in 50% of the wells (ID50) was calculated by the Reed-Muench formula (Reed and Muench, 1938).

### In vitro surrogate virus neutralising test (sVNT)

As an independent validation of the neutralising potential of sera from vaccinated or infected animals an FDA approved surrogate virus neutralising test (sVNT)^35^, manufactured by GeneScript, was applied. This test measures the ability of antibodies in the sera of vaccinated or infected animals to inhibit binding of horseradish peroxidase (HRP) conjugated recombinant WT SARS-CoV-2 RBD to plate-bound human-ACE2 protein. Results are expressed as percent inhibition.

### RBD Multiplex Binding Assay

A cocktail of RBD-variant coupled beads were incubated with serial dilutions of sera from vaccinated mice. Relative RBD antibody binding was detected using anti-mouse IgG R-Phycoerythrin (PE) Conjugate. The binding of IgG was detected as phycoerythrin-labelled reporter measured as MFI (Median Fluorescence Intensity). Results are expressed as half-maximal effective dilution (ED50). SARS-CoV-2 RBD variants were selected from the GISAID RBD surveillance repository.

### RBD-ACE2 multiplex inhibition assay

A cocktail of RBD-variant coupled beads were incubated with serial dilutions of sera from vaccinated mice. Samples were incubated for 1 hour before addition of Biotinylated human ACE2. After further 1hr incubation wells were washed and ACE2 binding was detected with Streptavidin followed by the addition of R-Phycoerythrin Biotin Conjugate. The binding of ACE2 was detected as phycoerythrin-labelled reporter measured as MFI (Median Fluorescence Intensity). Maximal ACE2 binding MFI was determined by the mean (quadruplicate) of ACE2 only (no inhibitor) controls. Results are expressed as half-maximal inhibitory dilution (ID50).

### Multiplex surrogate virus neutralization test (sVNT)

Multiplex sVNT against 20 different sarbecoviruses was performed as previously described^47^. Briefly, Luminex avidin beads coated with biotinylated RBD proteins were incubated with serial dilutions of sera from vaccinated mice for 15 min before addition of R-Phycoerythrin-conjugated human ACE2, followed by 15 min-incubation. The binding or loss of binding of ACE2 was measured as MFI by the Luminex MagPix instrument. Results are expressed as neutralization titer 50%.

### *In vitro* pseudovirus neutralisation test

To determine the neutralizing activity of hamster plasma, heat inactivated plasma was serially diluted and incubated with 200 TCID50 of pseudovirus expressing SARS-CoV-2 Spike of WT (HKU-13). The plasma-virus mixtures were added to HEK 293T-hACE2 cells. After 48 hours, infected cells were lysed, and luciferase activity was measured using Luciferase Assay System kits (Promega) in a Victor3-1420 Multilabel Counter (PerkinElmer). The 50% inhibitory dilution (ID50) of each plasma sample were calculated using non-linear regression in GraphPad Prism v8 to reflect anti-SARS-CoV-2 potency.

### In vitro plaque reduction neutralisation test

An in vitro plaque reduction neutralisation test (PRNT) measured the level of SARS-CoV-2-specific nAb in sera of immunised hamsters. Diluted sera samples were incubated with an equal volume of medium containing approximately 200 pfu/0.1 ml of SARS-CoV-2 WA-01/USA (WT) virus. The 90% neutralizing titres (PRNT90) were calculated as the highest dilution of serum showing >90% neutralisation of virus. The titre of non-neutralised virus was calculated as the mean of triplicate wells of virus incubated with diluent only, and that value was used to calculate the 90% neutralising titre.

### Mouse SARS-CoV-2 challenge model

Protective efficacy against upper (nasal turbinates) and lower (lung) airways infection was assessed using a mouse SARS-CoV-2 challenge model with a human clinical isolate of SARS-CoV-2, VIC2089 (N501Y) variant (hCoV-19/Australia/VIC2089/2020) or a naturally arisen beta (K417N, E484K, N501Y) variant B.1.351. Vaccinated and unvaccinated control mice were aerosol challenged with 1.5 × 10^7^ TCID50 infectious units of venturi-nebulised VIC2089 or B.1.351. Three days later challenged mice were euthanised and infectious virus titres (TCID50; 50% tissue culture infectious dose) in the lungs and nasal turbinates of individual mice were determined by titrating lung and nasal supernatants on Vero cell monolayers and measuring viral cytopathic effect (CPE) 5 days later.

### Hamster SARS-CoV-2 challenge models

Two separate hamster facilities were used for these studies. At The University of Hong Kong (HKU), protective efficacy against upper (nasal turbinates) and lower (lung) airways infection was assessed using a golden Syrian hamster (Mesocricetus auratus) SARS-CoV-2 challenge model with a human clinical isolate of WT SARS-CoV-2 (HKU-13 strain, GenBank accession number MT835140). The experiments were approved by the HKU Committee on the Use of Live Animals in Teaching and Research (CULATR). Vaccinated and unvaccinated control hamsters were challenged intranasally with 50µl containing 10^5^ plaque forming units (pfu) of virus under ketamine/xylazine anaesthesia^65^. Four days later, the challenged hamsters were euthanised and infectious virus titres (pfu/ml) in the lungs and nasal turbinates of individual hamsters were determined by titrating lung and nasal supernatants on Vero-E6 cell monolayers and counting plaques 3 days later.

At Colorado State University, protective efficacy against upper (oropharynx) and lower (lung) airways infection was assessed using a golden Syrian hamster SARS-CoV-2 challenge model with a human clinical isolate of SARS-CoV-2, WT (WA-01/USA) or a naturally arising Beta (K417N, E484K, N501Y) variant, B.1.351. These studies were conducted under approval number 1106 from the Colorado State University Institutional Animal Care and Use Committee (13Jul2020). Hamsters were vaccinated with 30, 10 or 3µg of the WT RBD-hFc dimer with MF59 or Beta RBD-hFc dimer with MF59, receiving 2 intramuscular doses on days 0 and 21. A 3rd dose was administered on day 63, and challenge performed on day 85. Hamsters were challenged under ketamine-xylazine anesthesia by intranasal instillation of 1 × 10^4^ pfu of SARS-CoV-2 virus in a volume of 100µl. Oropharygeal swabs were collected on days 1, 2, and 3 post-challenge. Three days post-challenge, hamsters were sacrificed and infectious virus titres (pfu/gram) in turbinate, cranial lung and caudal lung tissue were determined by plaque assay of homogenates on Vero cell monolayers. Neutralising antibody titres against the WT strain were assessed in sera collected 2 weeks after the third dose of vaccine using the plaque reduction neutralisation test (PRNT) using a 90% cutoff.

## Data Availability

All data produced in the present study are available upon reasonable request to the authors

## Funding and Acknowledgements

The authors are grateful to Professor Heidi Drummer, and Dr Rob Center, Burnet Institute for provision of the beta RBD monomer for ELISA studies. This work was supported by grants from the Medical Research Future Fund (MRFF) Awards (2005544, 2002073, 2002132), The Jack Ma Foundation, and National Health and Medical Research Council of Australia (NHMRC; 1113293, 2002317 and 1116530). GD was supported by philanthropic funds from IFM, DIG was supported by an NHMRC Senior Principal Research Fellowship (1117766) and an NHMRC Investigator Award (2008913). KK was supported by the NHMRC Leadership Investigator Grant (1173871), KS also received support from the A2 Milk Company. The Melbourne WHO Collaborating Centre for Reference and Research on Influenza is supported by the Australian Government Department of Health. The work at Duke-NUS is supported by grant from Singapore National Medical Research Council (MOH-COVID19RF-003). The authors also gratefully acknowledge the support of many others including: Rebecca Plavcak, Gabi Panoschi and The Doherty Institute BioResources Facility staff; Pin-Shie Quah and Martin Elhay (Doherty Institute); Judith Scoble, John Power, Pat Pilling, Susie Nilsson, Tram Phan (CSIRO); Drew Brockman and Peter Tapley (Agilex Biolabs); Kym Hoger, Karen Hughes, Ben Hughes, Paul Young and Keith Chappell (The University of Queensland), Jillian Bennet (Tanawell Consulting), Lena Miloradovic, Gary Grohman and Patricia Vietheer (Biointelect). The authors also acknowledge the facilities, and the scientific and technical assistance of the National Biologics Facility (NBF), The University of Queensland. NBF is supported by Therapeutic Innovation Australia (TIA). TIA is supported by the Australian Government through the National Collaborative Research Infrastructure Strategy (NCRIS) program.

## Author contributions

GD and CYW performed most of the mouse experiments. NAG designed and produced the initial vaccines. JPC, KD performed mouse virus challenge experiments. RZ, JFWC, ZWC, AEH, RB performed hamster experiments. GD, CYW, SLG, SR, PE, FLM, TS, MG, EL, JM, CWT, JW, WZ, ML, JMM, EV, JT, KK, CWP, TG, LW, SJK, AKW, SRL, KS, AC, MP, TM performed assays, provided samples or reagents and/or advised or directed experimental work. GD, NAG, SR, TN, DCJ, DFJP, DIG conceived the study, and analysed the data. DIG and GD wrote the paper with input from all authors.

## Figure Legends

**Supplementary Figure 1:**
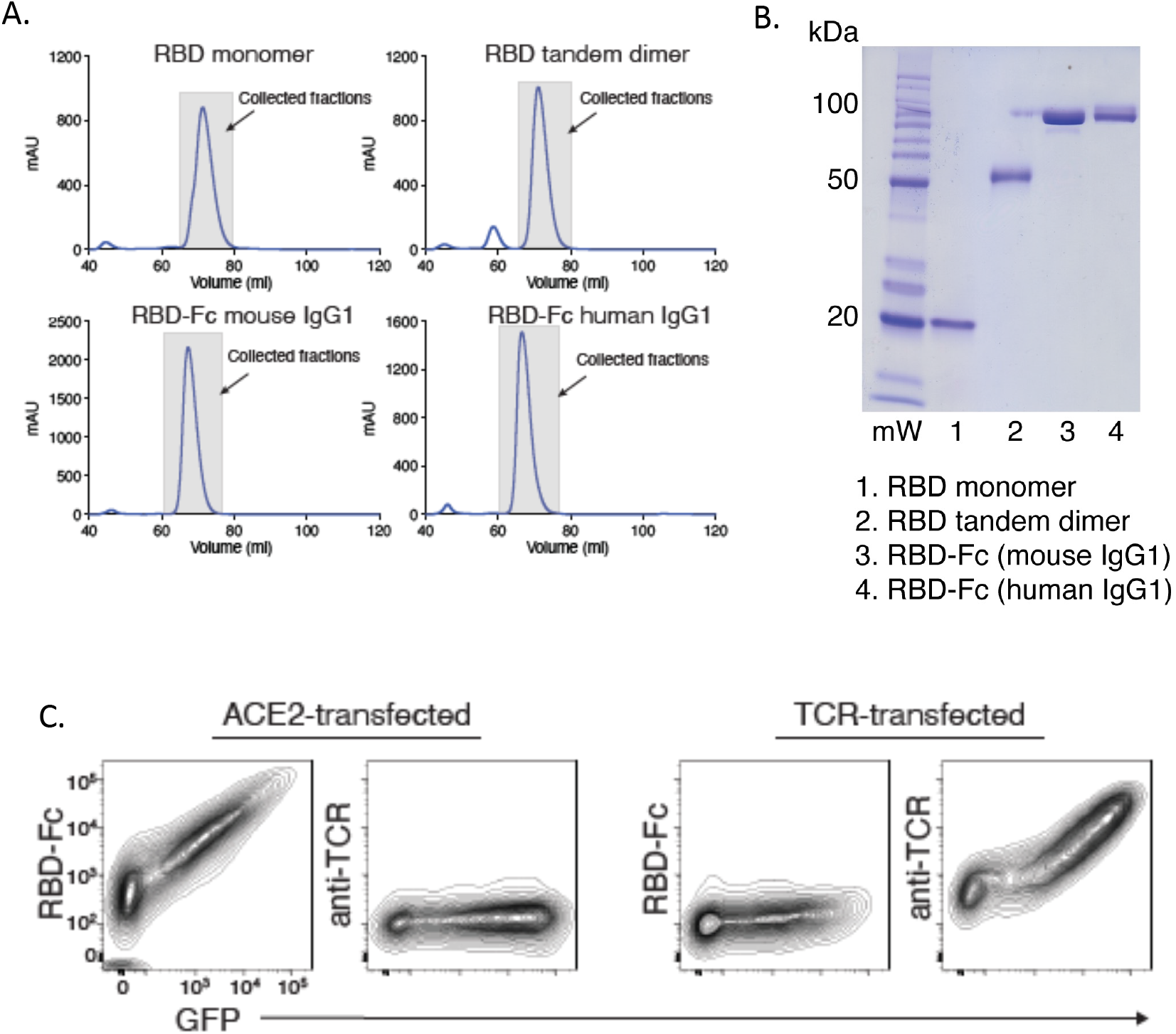
Generation of WT RBD proteins. **(A)** FPLC elution profiles from gel filtration size exclusion chromatography. Collected fractions are indicated. RBD monomer was eluted from a Superdex-75 column whereas RBD dimer and the RBD-Fc proteins were eluted from Superdex-200 column. **(B)** SDS-PAGE gel of the four WT RBD proteins post gel-filtration. **(C)** Flow cytometry contour plots showing HEK293T cells transiently transfected to express human ACE2, or an irrelevant protein (a T cell receptor (TCR)) as a negative control, stained with RBD-Fc (mouse IgG1) or anti-TCR antibodies.

**Supplementary Figure 2:**
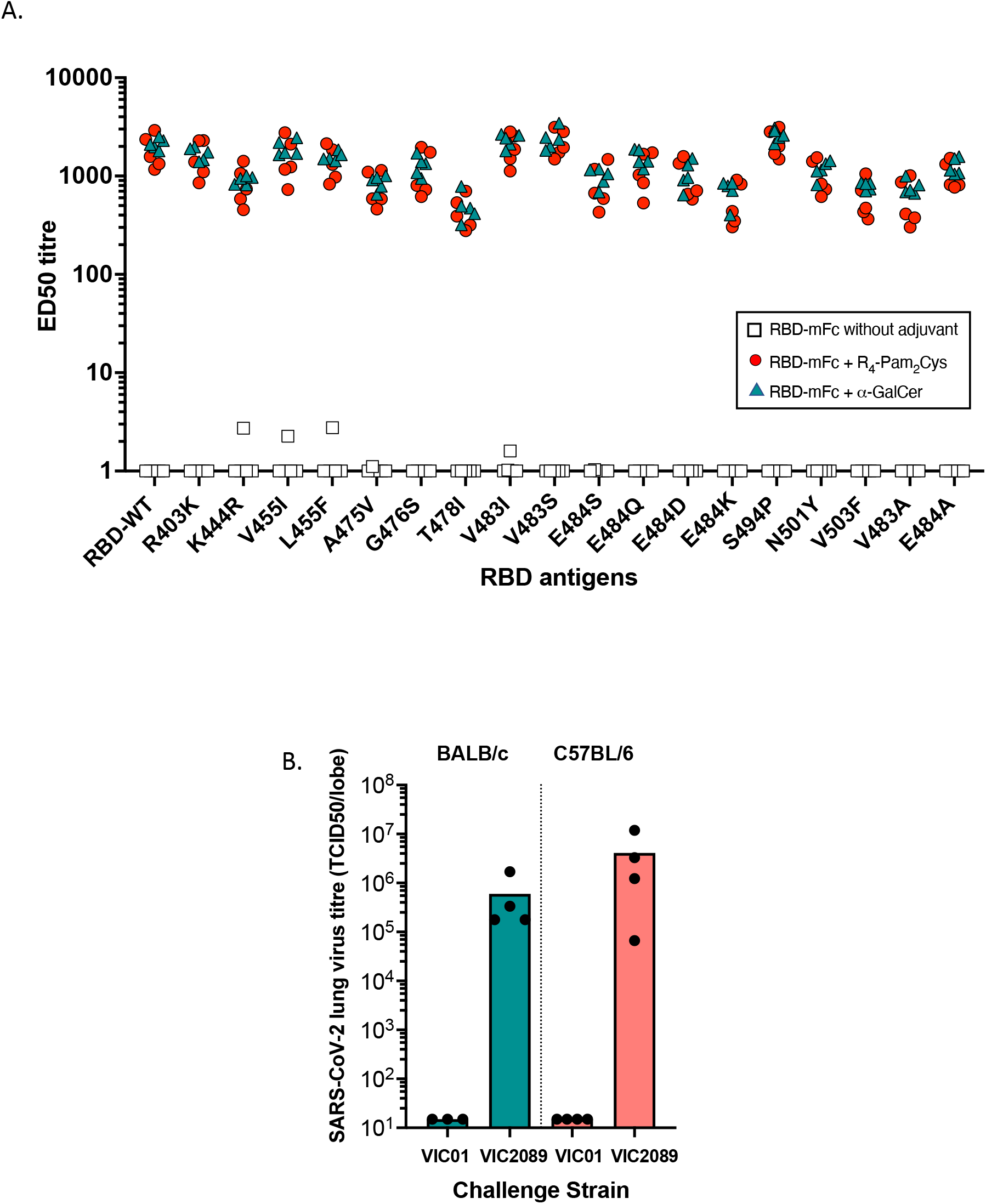
(A) IgG antibody responses to WT and mutant RBDs in mice vaccinated subcutaneously on day 0 and 29 with 10µg WT RBD-mFc in the presence or absence of adjuvant. Multiplex bead-based assay was used to assess IgG antibody responses to WT and mutant RBDs in secondary sera collected on day 41. Half-maximal effective dilution (ED50) is indicated for mice vaccinated with WT RBD-mFc in the absence of adjuvant (clear squares), or WT RBD-mFc administered with 0.3 nmole R4-Pam2Cys (grey circles) or with 0.2µg α-GalCer (black triangles). **(B)** Productive infection of mice with a naturally arising D614G/N501Y SARS-CoV-2 clinical variant (VIC2089). BALB/c or C57BL/6 mice were aerosol challenged with 1.5 × 10^7^ TCID50 infectious units of venturi-nebulised VIC01 (WT) or VIC2089 (N501Y) variant. Three days later mice were euthanised and infectious virus titres (TCID50) in the lungs of individual mice were determined by titrating homogenised lung supernatants on Vero cell monolayers and measuring viral CPE 5 days later. Bars depict mean results.

**Supplementary Figure 3:**
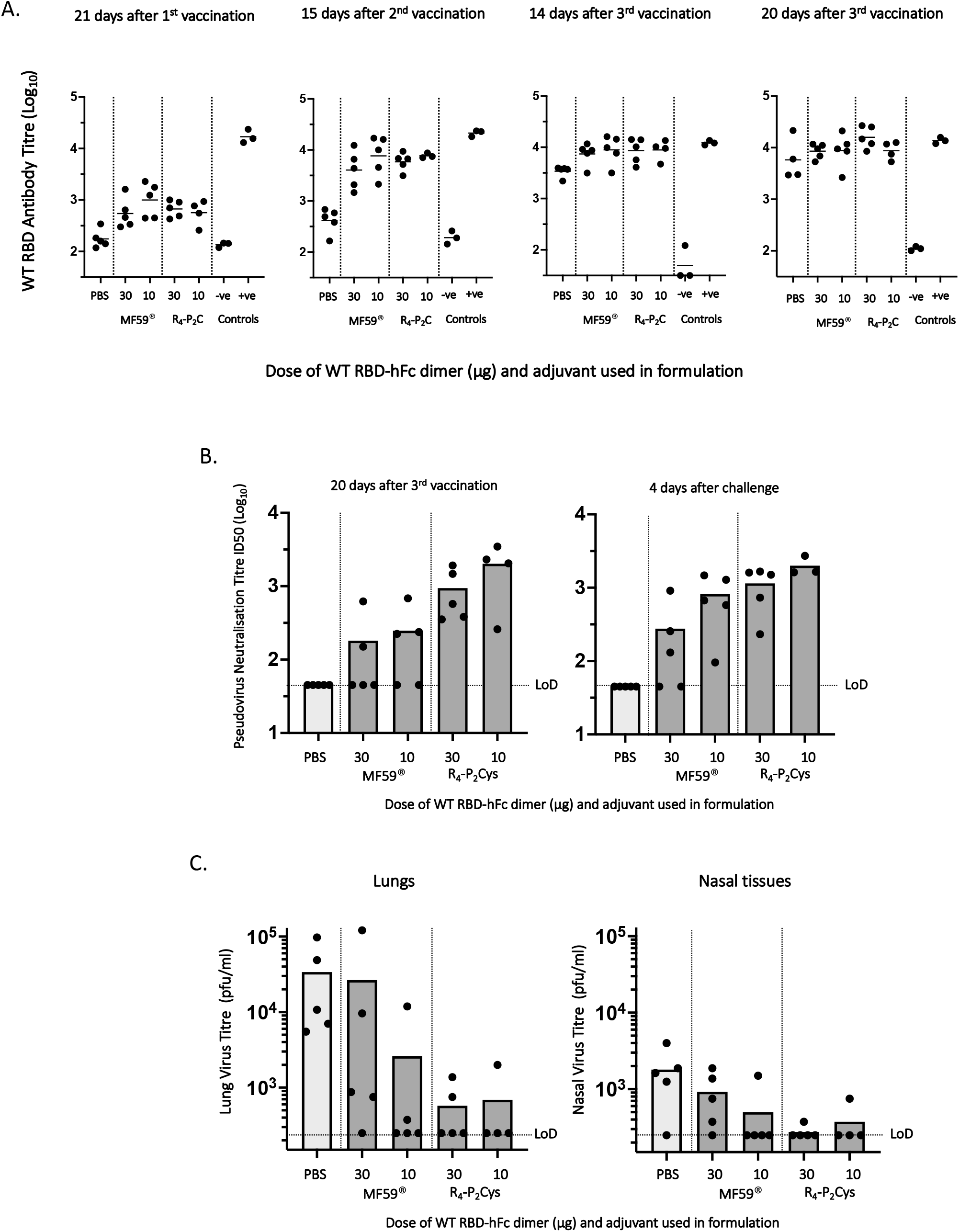
Immunogenicity and protection in hamsters using SARS-CoV-2 WT RBD-hFc vaccine. Hamsters at the University of Hong Kong were vaccinated intramuscularly on days 0, 21 and 49 with 30 or 10µg of WT RBD-hFc formulated with either MF59® or R4-Pam2Cys. On day 72 (23 days after the third vaccination) hamsters were challenged with SARS-CoV-2 virus (WT HKU-13 strain). **(A)** ELISA data showing anti-RBD antibody titres from individual hamster plasma samples at each timepoint. -ve control = normal hamster plasma and +ve control = day 21 plasma from a hamster vaccinated with an adenovirus vector expressing S of WT HKU-13 strain. **(B)** Pseudovirus neutralisation assay using a pseudovirus expressing SARS-CoV-2 Spike of the WT HKU-13 strain was used to assess nAb in serum samples collected 20 days after the third vaccination, and 4 days after the hamsters were challenged. **(C)** On day 76 (4 days after challenge) the hamsters were euthanised and the viral load in lungs and nasal tissues was determined by plaque assay using Vero-E6 cell monolayers. Titres are expressed as plaque forming units (pfu)/ml. Mean results are depicted with lines (A) or bars (B and C).

**Supplementary Figure 4:**
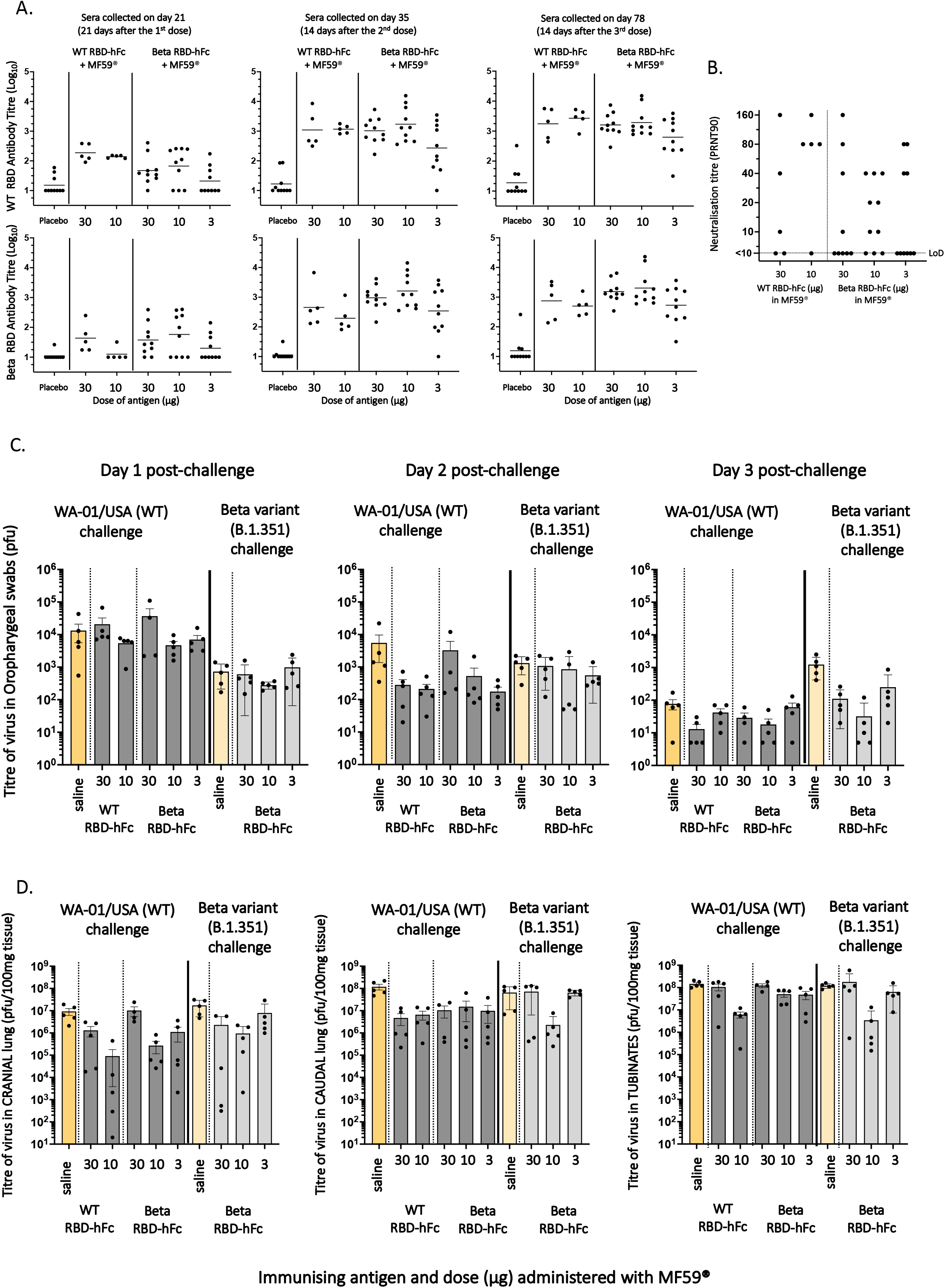
Immunogenicity and protection in hamsters using SARS-CoV-2 WT RBD-hFc and beta variant RBD-hFc vaccines. Hamsters at Colorado State University were vaccinated intramuscularly on days on days 0, 21 and 64 with 30 or 10µg of WT RBD-hFc + MF59® or with 30, 10 or 3µg of beta RBD-hFc + MF59®. On day 85 (21 days after the third vaccination) hamsters were challenged with either WA-01/USA (WT) or beta variant (B.1.351). **(A)** WT RBD and beta RBD-specific antibody titres in sera of hamsters bled 21 days after the first vaccination or 14 days after the second and third vaccinations. **(B)** nAb titres in sera collected 14 days after the third vaccination against the SARS-CoV-2 WT strain (WA-01/USA) were determined using the plaque reduction neutralisation test (PRNT). The 90% neutralizing titre (PRNT90) was calculated as the highest dilution of serum showing >90% neutralisation of virus. **(C)** Oropharygeal swabs were collected daily from each hamster day 1 to day 3 after challenge and prior to the animals being euthanised, and viral load determined by plaque assay using Vero cell monolayers (pfu/swab). **(D)** Viral load in cranial and caudal lung tissue, and turbinate tissue from each hamster on day 3 post-challenge were determined by plaque assay using Vero cell monolayers (pfu/100mg tissue). LoD: Limit of detection. Mean results are depicted with lines (A) or bars (C and D). Error bars depict SEM.

**Supplementary Figure 5:**
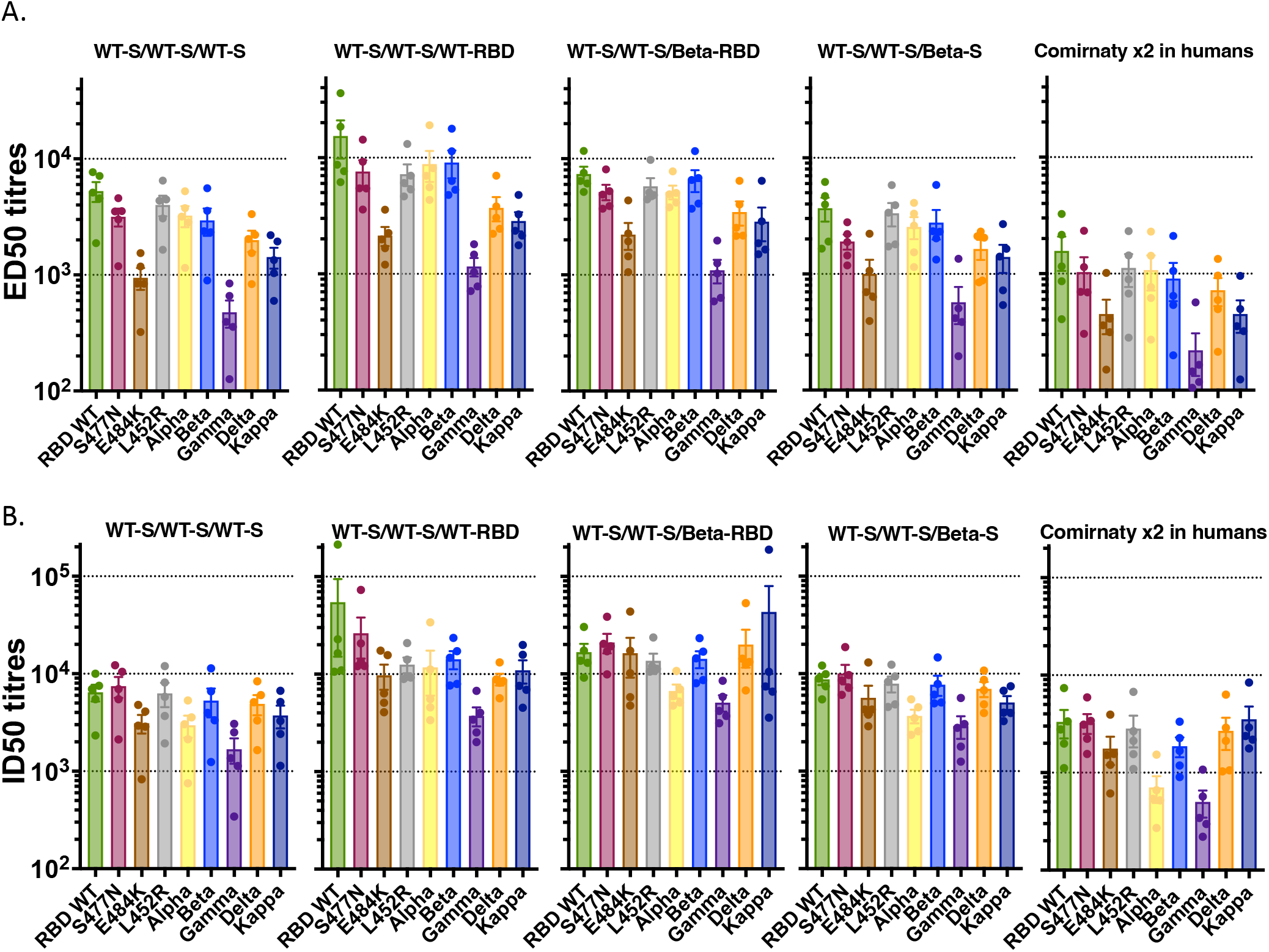
Immunogenicity of heterologous third dose boosting of mice previously vaccinated with 2 doses of WT spike vaccine. C57BL/6 mice (n=5) were vaccinated intramuscularly on days 0, 21 and 70 with 4.5µg of a spike antigen or 10µg of an RBD-hFc antigen in the presence of MF59®. The vaccine combinations administered, and the specific sequence of vaccine administration is as indicated above each graph. **(A)** Tertiary (day 86) sera collected 16 days after the third vaccination were assessed using the multiplex bead-based assay for their ability to bind to WT RBD and 8 RBD variants. Half-maximal effective dilution (ED50) is indicated for each serum sample. **(B)** The day 86 sera were also assessed using the RBD-ACE2 multiplex inhibition assay for their ability to inhibit the binding of ACE2 to WT RBD and the 8 RBD variants. Half-maximal Inhibitory dilution (ID50) is indicated for each serum sample. ED50 and ID50 levels measured in sera from human vaccinees given 2 doses of *Comirnaty*^*TM*^ (Pfizer mRNA S vaccine) on day 0 and 21 and bled on day 35 are also displayed in the right-hand graphs in A and B. Bars depict means and SEMs.

**Supplementary Figure 6:**
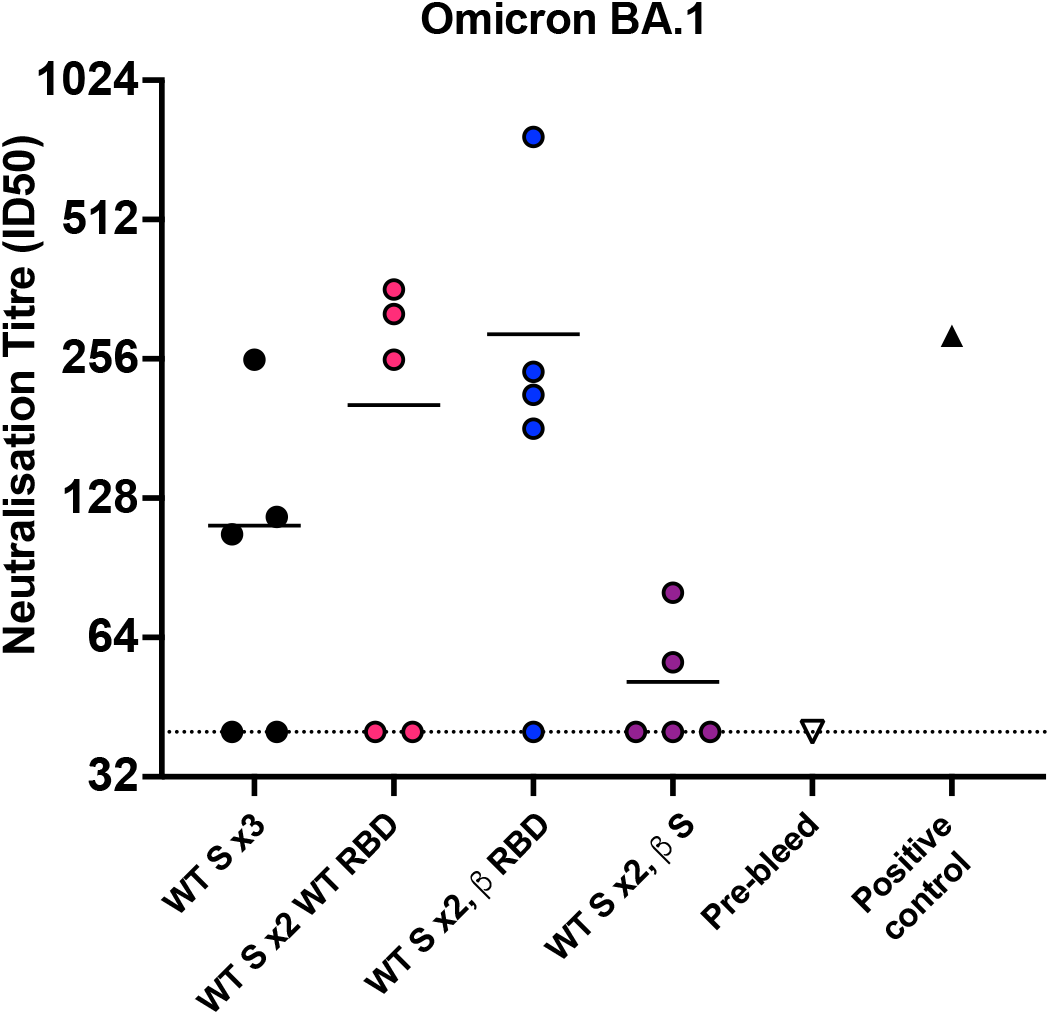
Serum samples taken 50 days following the third immunisation, from C57BL/6 mice vaccinated intramuscularly on day 0 and 21 with 4.5µg of the WT-spike with MF59® and boosted as described in the figure, were tested in an omicron-BA.1 variant microneutralisation assay using TMPRSS2 overexpressing Vero cells. In this assay, 10 TCID50 of virus per well was used to improve sensitivity. Data are average of duplicates. Horizontal lines depict means.

